# Leveraging nationwide health care records in Estonia to identify the genetic background of understudied disease phenotypes

**DOI:** 10.1101/2025.03.18.25324091

**Authors:** Erik Abner, Anastasiia Alekseienko, Kanwal Batool, Klavs Jermakovs, Mihkel Jesse, Hele Haapaniemi, Satu Strausz, Lehte Türk, Sviatoslav-Oleh Savchak, Estonian Biobank Research Team, Health Informatics Research Team, FinnGen, Anu Reigo, Teele Palumaa, Hanna M. Ollila, Jaanika Kronberg, Urmo Võsa, Tõnu Esko, Priit Palta

## Abstract

Nationwide health records linked to population biobanks can expand genetic discovery into clinical phenotypes that are poorly captured in hospital-centred datasets. We performed genome-wide association analyses of 5,491 ICD-10-based disease phenotypes in 206,159 Estonian Biobank participants using imputed genotype data across 18.8 million single-nucleotide and insertion-deletion variants.

Across the disease phenome, we identified 3,222 genome-wide significant loci, with strongest added value for outpatient-enriched, recurrent, and earlier-onset conditions. Fine-mapping prioritised candidate causal variants across study-wide significant loci, while coding variant analyses identified 754 protein-altering variant-trait associations outside the HLA region, including high-confidence signals in various dermatological, anaemia, congenital, and metabolic traits. A separate HLA analysis identified 744 HLA-trait associations across infectious, autoimmune, and skin-related phenotypes.

As an example of discovery in an understudied phenotype, we highlight pityriasis versicolor, a superficial fungal infection with 34 loci in EstBB-FinnGen meta-analysis, including a rare Estonian-enriched splice-disrupting *TNFSF15* variant. These results establish EstBB as a resource for mapping genetic architecture across clinically diverse and underrepresented disease phenotypes.

## Introduction

Genome-wide association studies (GWAS) have significantly advanced understanding of the genetic basis of complex human diseases and traits by enabling systematic identification of genetic variants associated with diverse phenotypes across various populations^1,2^. GWAS has led to numerous breakthroughs by enabling the development of more personalised medical predictions and interventions via polygenic risk scores (PRS), as well as clarification of the disease mechanisms^3,4^.

As sample size is critical in GWA studies, larger study cohorts significantly enhance statistical power to detect disease-associated variants. Some of the most extensive GWA studies focusing on single disease traits include type 2 diabetes, involving approximately 1.4 million individuals^5^, and major depression with over 1.2 million participants^6^, while quantitative traits like height have encompassed 5.4 million samples^7^. Although large single-trait GWAS provide valuable insights into the genetic architecture of these traits, comprehensive analyses across multiple traits can facilitate the identification of shared genetic backgrounds and comorbidity patterns.

During the last decade, numerous large-scale population-based collections have emerged globally, offering extensive datasets from thousands to millions of individuals, typically characterised through detailed questionnaires, comprehensive laboratory assessments, and electronic health record (EHR) linking, without selecting participants based on specific diseases^8^. Among the most prominent are the UK Biobank^9^ and the FinnGen project^10^. Several initiatives are focusing on diverse and non-European populations, including the China Kadoorie Biobank, BioBank Japan, the Mexican Biobank Project, and the African BioGenome Project^11–14^. The Estonian Biobank (EstBB) includes approximately 20% of Estonia’s adult population and integrates comprehensive linking to EHRs, facilitating detailed analyses of diagnostic profiles^15,16^.

Despite these efforts, large-scale biobanks often face limitations, including challenges in harmonising phenotypes, reliance on self-reported data prone to recall bias and other inaccuracies, and underrepresentation of primary-care and deeply characterised phenotypes^17^. These limitations restrict the scope of genetic analyses, particularly when investigating complex traits that involve heterogeneous subtypes or endo-phenotypes^18^. With nationwide electronic health records in place since 2004 and ICD-10 used consistently across care settings, EstBB addresses these challenges through linkage to standardized medical registries and continuous refinement of high-quality phenotypes using multiple complementary data sources^15^. Additionally, around 95% of ICD-10-based diagnoses in EstBB originate from outpatient and primary care settings, enabling in-depth analyses of conditions captured beyond inpatient care, earlier-stage disease, and phenotypes that are often underrepresented in hospital-based datasets^16^.

Here, we analysed 5,491 ICD-10-based traits and disease phenotypes in the Estonian Biobank, creating a large-scale genetic association resource based on nationwide health records and population-specific imputation. The atlas includes 3,222 genome-wide significant loci, hundreds of protein-altering variant associations, and a broad map of HLA allele-trait associations across clinical disease categories. Through fine-mapping, cross-biobank comparisons, coding variant prioritisation, and phenotype-specific follow-up, we show how outpatient-rich population biobanks can extend genetic discovery into common, recurrent, and previously underpowered clinical phenotypes. As an example, we use pityriasis versicolor to illustrate how such data can reveal host-genetic architecture for an understudied disease endpoint.

## Results

### Overview of the Estonian Biobank phenotypic data

The Estonian Biobank is linking health information annually from multiple national sources, with the Estonian Health Insurance Fund as the primary contributor^16^. The vast majority of diagnoses in the EstBB dataset originate from nationwide medical records (**Figure 1A**), with questionnaire-based self-reported diagnoses contributing to only 0.80% of all entries. The wealth of clinically verified information supports a reliable view of disease occurrence and longitudinal progression, whereas the age distribution of biobank participants (median age 52 years; median ICD-10 major category count 51) provides adequate coverage of diagnoses and follow-up time accumulated since early adulthood (**Figure 1B**).

**Figure 1.**
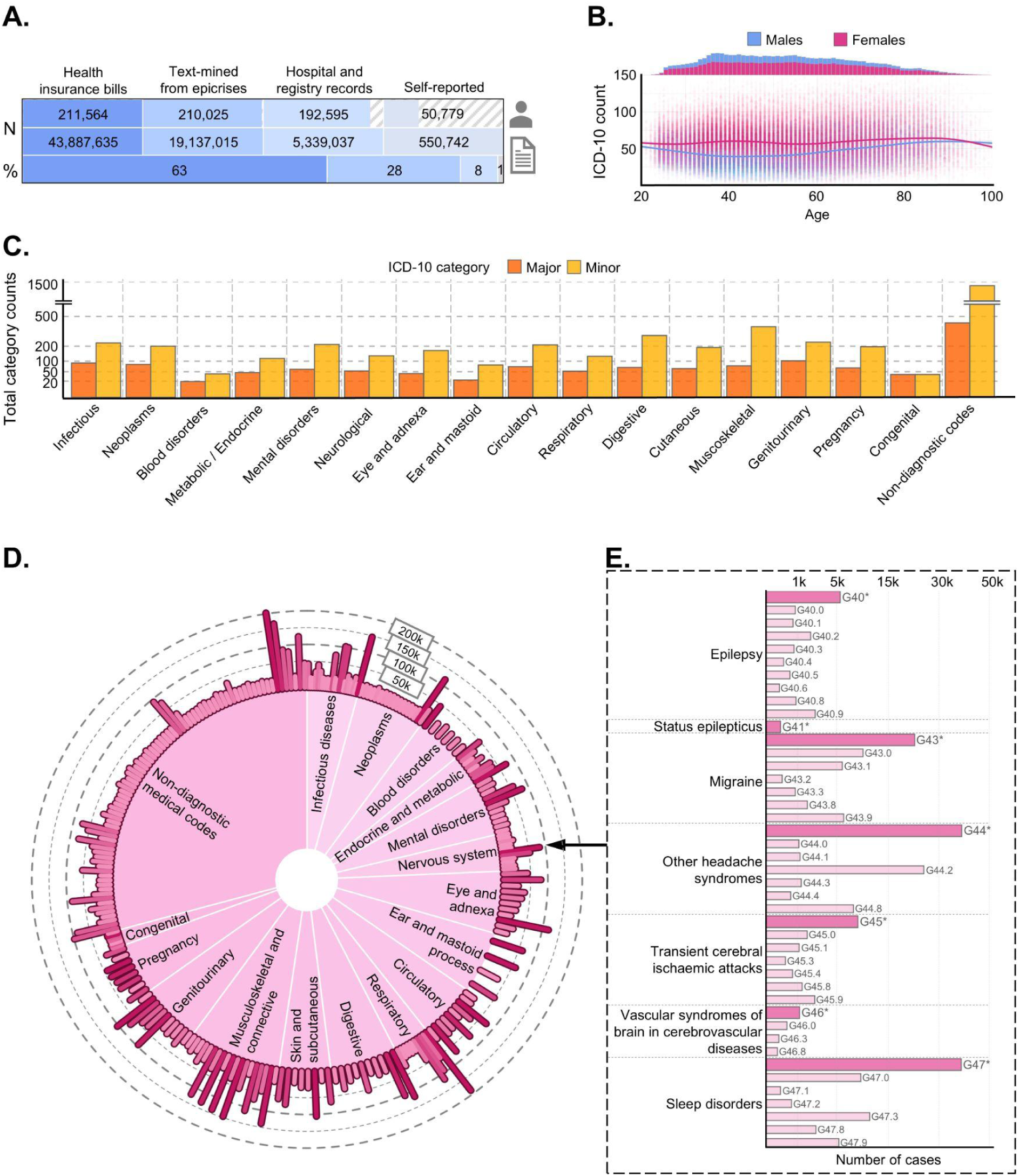
Overview of Estonian Biobank phenotypic data. **A** Distribution of health record sources in Estonian Biobank (2026 dataset). Bars show the number of individuals, ICD-10-based medical events, and the proportion of records contributed by each source; bars are stacked to 100%, hatched regions denote the remaining proportions of participants without the data from corresponding sources. **B** Age and sex distribution of Estonian Biobank participants included in the 2026 dataset, shown together with the number of unique ICD-10 major category diagnoses recorded per participant across age. The upper panel histogram shows the age distribution separately for males (blue) and females (purple), and the main panel scatterplot shows the distribution of ICD-10 counts by age and sex, with smoothed trend lines indicating the median ICD-10 counts. ICD-10-based events were counted only once per participant for each code, so repeated registrations of the same diagnosis do not inflate disease counts. **C** Counts of ICD-10-based traits included in the GWAS analyses, with at least hundred participants, grouped by broad diagnostic criteria. Major category codes are shown in orange, and minor subcodes in yellow. **D** Case counts of ICD-10 diagnoses in primary diagnostic groups. The colour gradient indicates the number of cases per group. Dashed horizontal lines indicate thresholds of 50,000, 100,000, 150,000, and 200,000 cases. **E** Detailed view of the ICD-10 diagnostic group episodic and paroxysmal disorders (ICD-10 G40-G47), showing case counts of its major diagnostic categories and minor subcategories.

The phenotypes defined for this study included a total of 5,491 medical ICD-10-based phenotypes: 4,884 universal, 545 female-specific, and 62 male-specific (**Methods**). Of those, 1,350 correspond to major diagnostic categories (such as E11 - “Type 2 diabetes mellitus”) and 4,141 represent minor subcategories (e. g. E11.0 - “Type 2 diabetes mellitus with coma”). The more granular subcodes capture clinically relevant differences in aetiology, anatomical site, severity, or complications, which can facilitate more specific phenotype resolution for genetic analysis (**Figure 1C; Supplementary Table 1**). Outpatient care-influenced disease categories were highly prevalent in the cohort, resulting in a high representation of infectious diseases, skin and subcutaneous tissue disorders, and diseases of the nervous system (**Figure 1D**). These traits are especially well captured for young adult biobank participants from outpatient records. This can be exemplified with the medical event occurrences in the episodic and paroxysmal disorders group (ICD-10 block G40-G47), which shows a strong enrichment due to common outpatient diagnoses including headaches, migraines, and sleep disturbances (**Figure 1E**). In contrast, conditions that are less commonly recorded in outpatient settings show lower prevalence in the dataset, exemplified by phenotype groups such as congenital anomalies (ICD-10 category Q*) (**Supplementary Table 2**).

### Genetic discovery across disease phenotypes in EstBB

To carry out the main GWAS analyses, we used imputed genotype data for 206,159 EstBB participants (see **Methods**), comprising of approximately 17.24 million single-nucleotide variants (SNVs) and 1.56 million indel variants^19^ and including 6.56 million common variants (MAF > 0.05), 4.02 million low-frequency variants (0.005 < MAF ≤ 0.05), and 8.23 million rare variants (MAF ≤ 0.005), providing coverage across the full allele frequency spectrum (**Supplementary Figure 2**). We then applied a set of post-GWAS analyses to interpret these associations across complementary levels of disease biology. SNP-based heritability and cross-trait LDSC analyses were used to quantify genetic contribution and shared genetic architecture across ICD-10 traits, while comparisons with UKBB and FinnGen assessed phenotype coverage, locus discovery, and cross-biobank genetic concordance. Study-wide significant loci were fine-mapped with SuSiE to prioritise credible sets and high-confidence candidate variants. In parallel, coding variant annotation, HLA allele association testing, and phenotype-specific follow-up analyses were used to identify interpretable protein-altering, immune-mediated, and population-enriched signals for downstream biological interpretation.

### SNP-based heritability and locus discovery

To quantify the genetic contribution to disease variation across the EstBB phenome, we estimated the SNP-based heritability for all studied phenotypes. Out of all 5,491 phenotypes, 1,958 (35.66%) had effective sample size higher than 4,500, and 297 phenotypes (5.41%) displayed a statistically significant SNP-based heritability after Bonferroni correction (*P* < 9.11 × 10^-6^), with a mean heritability (h^2^) of 0.066 among significant traits (**Supplementary Table 3**). Traits with the highest SNP-heritability included obesity (ICD-10 E66, h² = 0.224, *P* = 2.0 × 10^-75^), other specified hypothyroidism (ICD-10 E03.8, h² = 0.176, *P* = 1.63 × 10^-19^), and myopia (ICD-10 H52.1, h² = 0.175, *P* = 3.05 × 10^-71^). In contrast, infectious disease traits such as herpesviral infections (ICD-10 B00) and COVID-19 (ICD-10 U07) displayed considerably lower heritability estimates (h² = 0.036, *P* = 1.6 × 10^-7^ and h² = 0.034, *P* = 1.5 × 10^-13^, respectively), consistent with a larger contribution from exposure and environmental factors (**Supplementary Figure 3**).

We next assessed whether these heritability patterns translated into locus discovery across the disease phenome. Across all GWASs, we identified 3,222 genome-wide significant loci corresponding to 1,358 phenotypes, of which 859 loci reached study-wide significance (*P* < 1.00 × 10^-11^). The largest number of independent loci mapped to endocrine and metabolic traits (ICD-10 category E*; 268 loci), followed by neoplasms (ICD-10 C*-D5*; 97 loci) and eye and adnexa traits (ICD-10 H0* - H5*; 82 loci) (**Supplementary Table 4**). These categories are consistent with traits that exhibit substantial genetic contribution, supported by the high SNP-based heritability estimates among endocrine and metabolic traits (**Supplementary Table 3**). At the individual-trait level, myopia (ICD-10 H52.1) had one of the highest locus count with 30 loci (**Figure 2A**), consistent with both its high SNP-based heritability and its extensive prior GWAS literature^20,21^.

**Figure 2.**
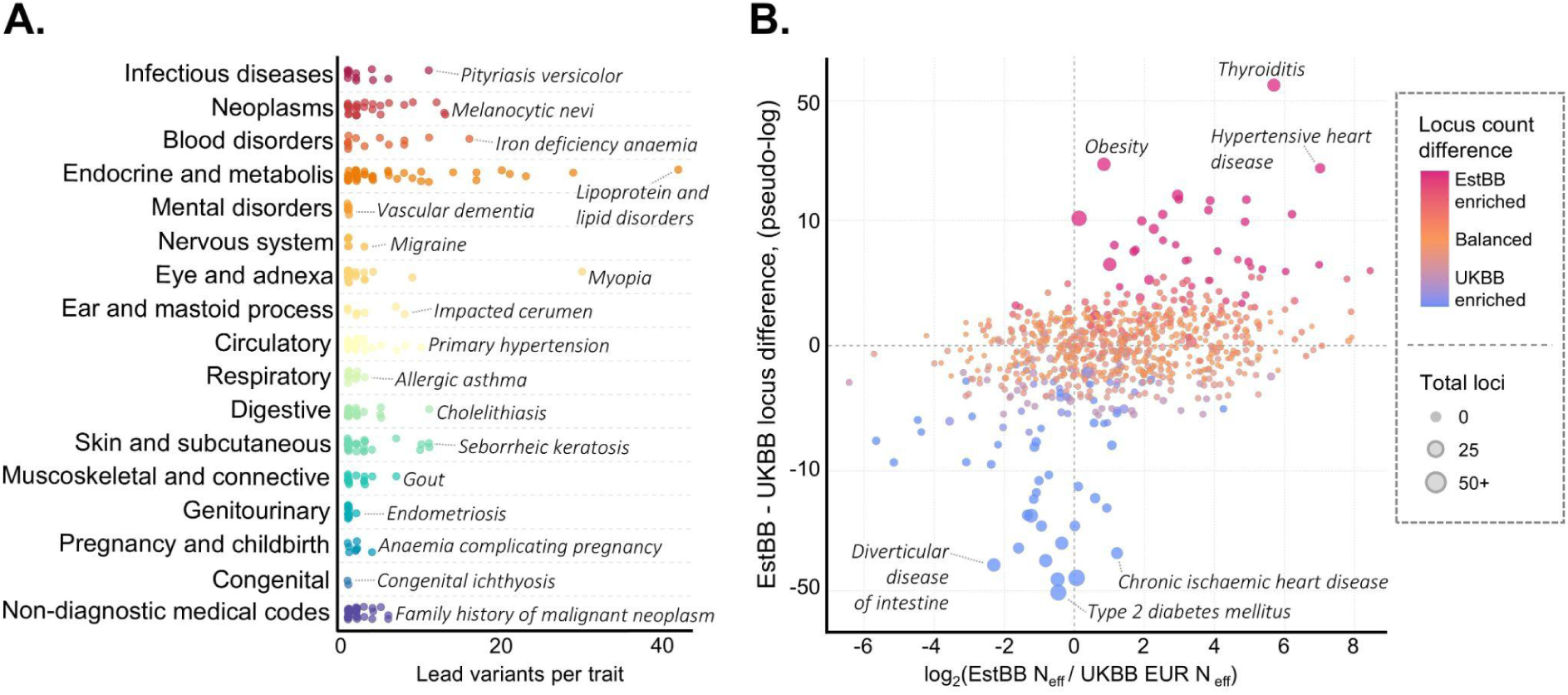
Genome-wide association discovery across ICD-10 traits in the Estonian Biobank. **A** Distribution of lead variants across ICD-10 disease categories. Each point represents one ICD-10 phenotype and shows the number of lead variants identified for that trait in EstBB. Phenotypes are grouped by ICD-10 category on the y-axis, and the x-axis shows number of lead variants per trait. Selected traits with particularly high numbers of lead variants are annotated. **B** Comparison of effective sample size and locus discovery between Estonian Biobank and UK Biobank (UKBB) across matched ICD-10 traits. Each point represents one ICD-10 phenotype. The x-axis shows the log_2_-transformed ratio of effective sample size in EstBB versus UKBB, with positive values indicating higher effective sample size in EstBB and negative values indicating higher effective sample size in UKBB. The y-axis shows the difference in the number of genome-wide significant loci between EstBB and UKBB, displayed on a pseudo-log axis to improve visualization of small locus-count differences while retaining larger positive and negative differences. Point colour reflects the EstBB minus UKBB locus difference. Point size indicates the total number of significant loci detected across both biobanks. The upper-right quadrant contains traits with higher EstBB power and more EstBB loci, while the upper-left quadrant highlights traits with more EstBB loci despite lower EstBB power. The three most EstBB-enriched and UKBB-enriched traits by locus difference are labelled.

### Outpatient-enriched phenotypes and comparison with UKBB

The high locus count for myopia illustrates how disease recording in EstBB differs from hospital-centred resources. Myopia was recorded in 51,585 EstBB participants (25.02% of the cohort), reflecting the systematic capture of common conditions through outpatient and primary-care records. Across the ICD-10 phenotypes, the dataset showed a median 15-fold excess of outpatient over inpatient events, and 91.19% of traits had at least two-fold outpatient enrichment (**Supplementary Table 2**). Together with the relatively even adult age distribution of the cohort (**Figure 1B**), this structure increases power for common, earlier-onset, recurrent, and non-hospitalised conditions.

This trait distribution particularity is also evident at the phenotype level. At the raw ICD-10 level, both resources contained cases for 11,004 codes, but EstBB had 20.88 million summed phenotype case counts compared with 4.91 million in UKBB, a 4.23-fold excess despite UKBB cohort being more than twice as large (**Supplementary Table 5**). Of the 4,884 non-sex-specific ICD-10 phenotypes defined in this study, 975 had no corresponding UKBB phenotype (**Supplementary Figure 1**). Among publicly available GWAS summary statistics for 921 UKBB traits, 771 traits matched our EstBB-based dataset (**Figure 2B, Supplementary Table 6**). UKBB captured more cases for late-onset conditions such as type 2 diabetes (ICD-10 E11; 16 EstBB loci versus 70 UKBB loci), diverticular intestine (ICD-10 K57; 2 versus 49 loci) and chronic ischaemic heart disease (ICD-10 I25; 2 versus 48 loci), consistent with its older recruitment profile^22^. In contrast, the younger, outpatient care-enriched EstBB cohort was better powered for earlier-onset conditions. In total, we detected study-wide significant loci in 131 EstBB traits where UKBB displayed no significant loci, with the largest differences observed for thyroiditis (ICD-10 E06; 43 loci), hypertensive heart disease (ICD-10 I11; 20 loci), and migraine (ICD-10 G43; 11 loci). The signal for hypertensive heart disease was particularly notable, as EstBB displays considerable case-enrichment (174-fold case-count enrichment), and 18 of the 20 EstBB loci were not significant in UKBB (*P* < 5 × 10^-8^), with the remaining two displaying only nominal evidence of association (*P* < 0.05). Two of the three strongest EstBB loci for this trait have also been reported in the GWAS Catalog for hypertensive heart disease (GWAS Catalog ID MONDO_0001302), including the known hypertension risk factor rs10857147 near *FGF5* gene^23^ and the obesity variant rs55872725 in intron 1 of *FTO*^24^ (**Supplementary Table 7**). Given that only nine loci for this trait have previously been reported in GWAS Catalog repository, these findings support the validity and added resolution of the EstBB signal.

The strongest EstBB case-count enrichments were concentrated in outpatient-captured and infection-related phenotypes, such as acute tonsillitis (ICD-10 J03; 7 versus 1 loci; 104-fold case-count enrichment) and influenza (ICD-10 J11; 3 versus 1 loci; 84-fold case-count enrichment). Traits with larger locus gains in EstBB also reflected healthcare-system and cohort differences, including obesity (ICD-10 E66; 31 versus 16 loci) and melanocytic naevi (ICD-10 D22; 17 versus 7 loci). Melanocytic naevi provided another example of a rather unique EstBB-specific locus discovery. Of the 17 EstBB loci for this trait, only three were genome-wide significant in UKBB (*P* < 5 × 10^-8^), while seven showed nominal evidence of association (*P* < 0.05). Notably, no genome-wide significant variants for melanocytic naevi are currently reported in the GWAS Catalog under ID MONDO_0005073.

### Genetic concordance with FinnGen

To assess genetic concordance with another external nationwide biobank resource, we compared EstBB GWAS results with publicly available FinnGen project GWAS summary statistics. EstBB and FinnGen represent genetically related Northern European population cohorts and are based on nationwide health registries with broadly comparable clinical coding practices^10^. We analysed 100 ICD-10 traits that met predefined inclusion criteria in both biobanks, had comparable clinical definitions, and were suitable for LD score regression (**Supplementary Table 8, Supplementary Figure 4**). Ninety-six traits showed nominally significant genetic correlations (*P* < 0.05), and 90 remained significant after Bonferroni correction (*P* < 5.00 × 10^-4^). Most traits that did not reach significance were immune-mediated phenotypes, including coeliac disease (ICD-10 K90.0), ankylosing spondylitis (ICD-10 M45), viral infections (ICD-10 A08), and other rheumatoid arthritis (ICD-10 M06). For these endpoints, a substantial fraction of genetic signal is likely driven by the HLA region, which was excluded from the cross-biobank LDSC analyses. Among the remaining traits, genetic correlations were consistently high (median rg = 0.832), supporting the robustness of EstBB phenotype definitions and their genetic concordance with an independent registry-based biobank.

Overall, these comparative findings with other biobank datasets position EstBB as a complementary resource for other genetic studies, adding power and phenotype coverage for clinically common and earlier-onset complex disease endpoints that are less well captured in older or more hospital-centred biobanks.

### Fine-mapping reveals shared genetic architecture

Overall, the GWAS identified 229,570 study-wide significant variant-trait associations (*P* < 1 × 10^−11^),, including 72,787 outside the HLA locus. To refine these association signals and prioritise likely causal variants within each locus, we performed Bayesian fine-mapping using SuSiE. This yielded at least one 99% credible set for 784 of 796 study-wide significant loci across 287 non-sex-specific traits, 60 of 62 independent study-wide significant loci across 26 female-specific traits, and one male-specific locus. Across all fine-mapped loci, the mean number of credible sets per locus was 1.38, and the mean credible set size was 22.37 variants. We defined high-confidence candidate variants as those with posterior inclusion probability (PIP) > 0.9, where PIP is the estimated probability that a variant is causal under the fine-mapping model, and identified such variants in 197 credible sets (16.88%) (**Figure 3A**). Among all credible sets, 511 (43.8%) contained at least one insertion-deletion (indel) variant, and in 15 loci an indel was the high-confidence top candidate. By contrast, 218 credible sets had an SNV as the high-confidence top candidate. In terms of allele frequency, credible sets collectively contained 11,984 common variants (MAF > 0.05) and 633 low-frequency variants (MAF ≤ 0.05), while high-confidence candidates comprised 64 common and 44 low-frequency variants (**Supplementary Figure 5, Supplementary Table 4**). Such credible sets and high-confidence candidates define a practical shortlist for downstream functional prioritisation.

**Figure 3.**
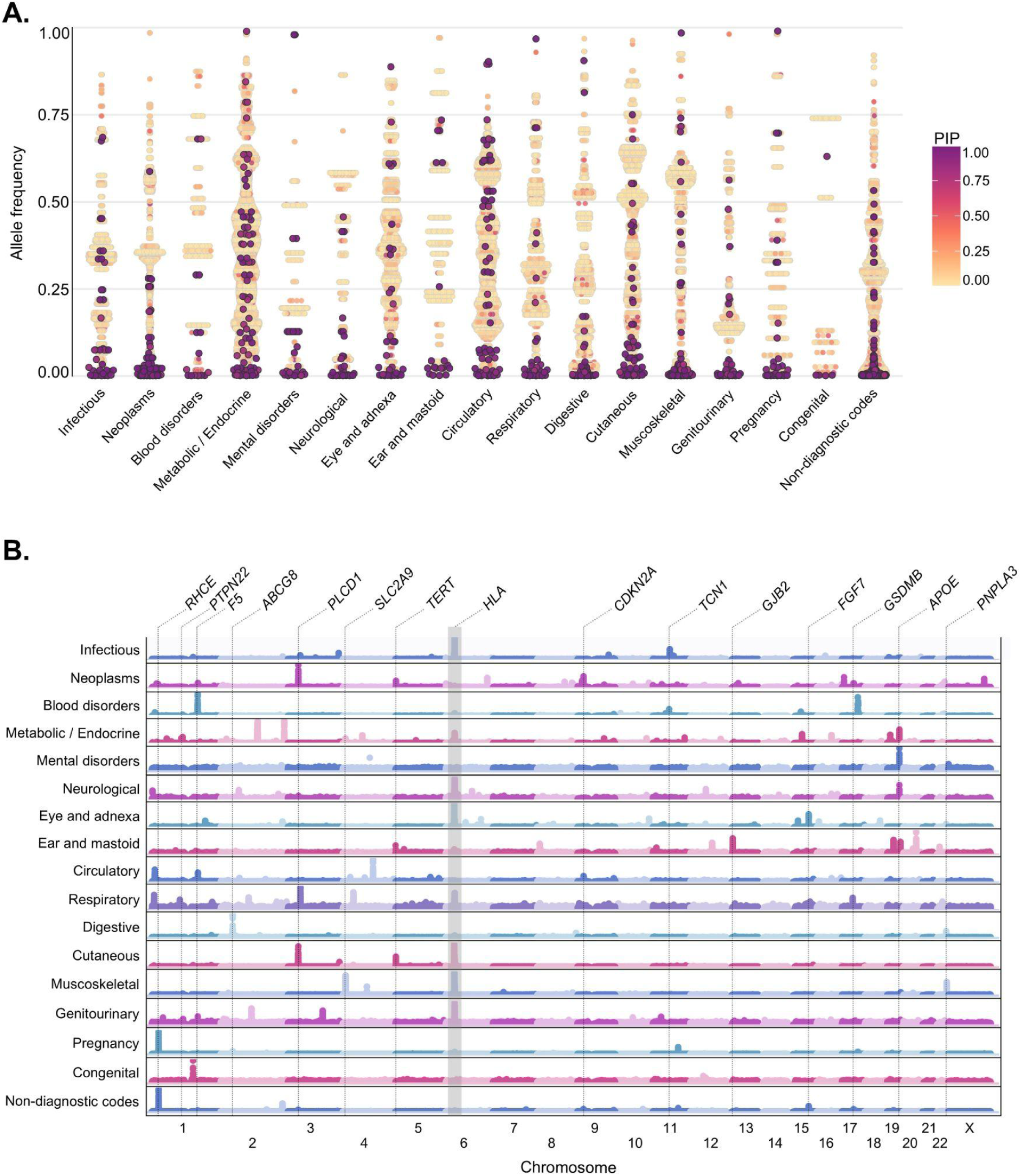
Landscape of significant and fine-mapped associations across ICD-10 disease categories. **A** Beehive plot of fine-mapped variants across ICD-10 disease categories. Each point represents a variant and is positioned by ICD-10 disease category on the x-axis and allele frequency on the y-axis. Points are displayed using jitter to reduce overlap. Variant colour reflects fine-mapping posterior inclusion probability (PIP). Variants with posterior probability <0.8 are shown as circles with a light gray outline, whereas variants with posterior probability ≥0.8 are shown as circles with a darker gray outline, highlighting higher-confidence signals. **B** Genome-wide distribution of pleiotropic associations across disease categories. The stacked scatterplot displays category-specific Manhattan plots for grouped disease traits. Each dot represents a genetic variant, plotted by chromosomal position on the x-axis and by the strongest association signal observed for that variant within the corresponding disease category on the y-axis as −log10(P). Selected pleiotropic or high-impact genes are annotated above the plots. Traits are grouped by broad disease categories, and chromosomes are shown in genomic order along the x-axis.

### Pleiotropic loci reveal shared disease architecture

Phenome-wide GWAS across many disease endpoints can identify loci that act across multiple clinical traits, either through shared disease biology or through effects on related diagnostic categories. We therefore assessed pleiotropy across study-wide significant loci, defining pleiotropic loci as those associated with more than five phenotypes. In total, eight genomic loci met this definition (**Figure 3B**). A noteworthy example was the chromosome 17p13.1 locus, which was associated with 12 dermatology-related phenotypes, including malignant skin neoplasms (ICD-10 C44), benign neoplasms (ICD-10 D18, D21, D23), follicular cysts (ICD-10 L72) and seborrhoeic keratosis (ICD-10 L82). In 11 of these 12 phenotypes, fine-mapping prioritised the same lead variant, rs78378222, a variant within the *TP53* gene. This allele is relatively common in EstBB (AF = 0.022) and showed its strongest effect among non-melanoma skin cancers, with the largest association observed for malignant neoplasm of skin of scalp and neck (ICD-10 C44.4; OR = 2.635; 95% CI = 1.977 - 3.511; *P* = 3.84 × 10^-11^). The allele is nearly two-fold enriched compared with non-Finnish European reference frequencies, increasing power to detect its effects in Estonia. Consistent with previous reports linking this variant to female reproductive traits, rs78378222 was also associated with leiomyoma of uterus among sex-specific endpoints (ICD-10 D25; OR = 1.626; 95% CI = 1.523 - 1.735; *P* = 1.08 × 10^-48^)^25,26^(**Supplementary Figure 6**).

*TP53* is central to genome integrity and tumour suppression^27^. The lead signal rs78378222 lies in the *TP53* 3′UTR (**Supplementary Figure 7**) and has previously been implicated in cancer susceptibility^28–30^, making it a plausible shared driver of the epithelial and dermatological phenotypes observed at this locus. Across these phenotypes, fine-mapping repeatedly converged on the same moderate-frequency variant, supporting a locus-specific pleiotropic effect rather than independent signals within *TP53*. We then asked whether this pattern simply reflected broader diagnostic similarity, and used inside-biobank LDSC to compare locus-level pleiotropy with genome-wide shared genetic architecture (**Supplementary Table 9**). Several phenotypes with the TP53 cluster showed strong genome-wide genetic correlation and marked record-level co-occurrence, as expected for closely related benign neoplasm and keratosis diagnoses (**Supplementary Figure 8, Supplementary Table 10**). However, the *TP53* signal also extended to phenotype pairs with weaker or non-significant genome-wide genetic correlation, including follicular cysts with seborrhoeic keratosis, malignant skin neoplasms, and haemangioma or lymphangioma. Thus, rs78378222 appears to mark a shared epithelial susceptibility mechanism that is not fully captured by overall genetic correlation or diagnostic co-occurrence between the traits.

### Dataset overlap supports established genetic signals

To assess whether EstBB revealed association signals absent from existing GWAS resources, we cross-referenced both lead and fine-mapped variants against the GWAS Catalog, Open Targets Genetics, and the Open Targets Platform databases. Most loci overlapped prior genetic evidence: 816 loci (94.99%) contained at least one credible-set variant reported in the GWAS Catalog, 814 (94.76%) in Open Targets Genetics, and 830 (96.62%) in the Open Targets Platform. An automated screening identified 18 candidate loci for which credible-set variants were absent from all three resources or had no reported significant associations. However, subsequent manual curation did not support presenting these as high-confidence novel biological findings, as several reflected partial overlap with known loci, mapping issues in the X-chromosome pseudoautosomal region, low imputation quality, or limited phenotype-specific interpretability (**Supplementary Table 11**). Therefore, as expected for a European-ancestry biobank of this size, EstBB did not yield high-confidence loci with no prior trait-level genetic evidence after manual curation. Instead, its main contribution lies in expanding the clinical contexts in which established and emerging genetic signals can be detected. This is particularly relevant for outpatient-enriched, recurrent, and earlier-stage diseases, where longitudinal health records can support meta-analyses and genetic studies of incident diagnoses, repeated events, and progression from early clinical presentation to severe disease.

### Coding variant associations across the EstBB

Population-specific biobanks provide a unique opportunity to discover coding variants with strong functional effects, many of which remain undetected in global datasets due to founder structure, drift or low allele frequencies elsewhere^31^. We therefore focused on coding variants with predicted protein-altering consequences and identified a total of 754 variant-trait associations outside the HLA locus (**Supplementary Table 12**).

Among these, 270 unique coding variants were associated with at least one phenotype and mapped to 184 genes, implicating 224 distinct traits (**Figure 4**). This included 79 unique variants and 252 variant-trait associations with CADD scores above 20, a threshold commonly used to mark variants among the most deleterious predicted substitutions^65^. Fine-mapping of the variant-trait associations yielded well-resolved credible sets for 290 associations, including 68 high-confidence variant-trait pairs (PIP ≥ 0.9). Among these, 20 were missense SNVs with clear protein-altering predictions and strong statistical support, providing immediately interpretable biological candidates. Notable examples included associations with dermatological phenotypes such as *PLCD1*:p.Ser460Leu with trichilemmal cysts (ICD-10 L72.1; OR = 3.858; 95% CI = 3.642 - 4.088; *P* = 8.0 × 10^-479^), *SCGB1D2*:p.Pro53Leu with Lyme disease (ICD-10 A69.2; OR = 1.194; 95% CI = 1.169 - 1.220; *P* = 5.33 × 10^−58^), *TYR*:p.Arg402Gln with malignant skin neoplasms (ICD-10 C44; OR = 1.194; 95% CI = 1.145 - 1.245; *P* = 1.12 × 10^−16^), *TLR1*:p.Ser602Ile with other rosacea (ICD-10 L71.8; OR = 0.825; 95% CI = 0.784 - 0.860; *P* = 1.41 × 10^−16^) and *OCA2*:p.Ala481Thr with actinic keratosis (ICD-10 L57.0; OR = 1.891; 95% CI = 1.605 - 2.227; *P* = 2.4 × 10^−14^). High-confidence coding signals also highlighted anaemia and metabolic traits, including *MTHFR*:p.Ala222Val with folate deficiency anaemia (ICD-10 D52; OR = 1.436; 95% CI = 1.339 - 1.540; *P* = 4.39 × 10^−24^), *F5*:p.Arg534Gln with coagulation defects (ICD-10 D68; OR = 5.202; 95% CI = 4.432 - 6.106; *P* = 1.61 × 10^−90^), multiple vitamin B12 deficiency anaemia associations mapping to vitamin B12 transporting genes *TCN1* and *TCN2*, and established lipid biology at *APOE* and *PCSK9* (**Supplementary Figure 9**).

**Figure 4.**
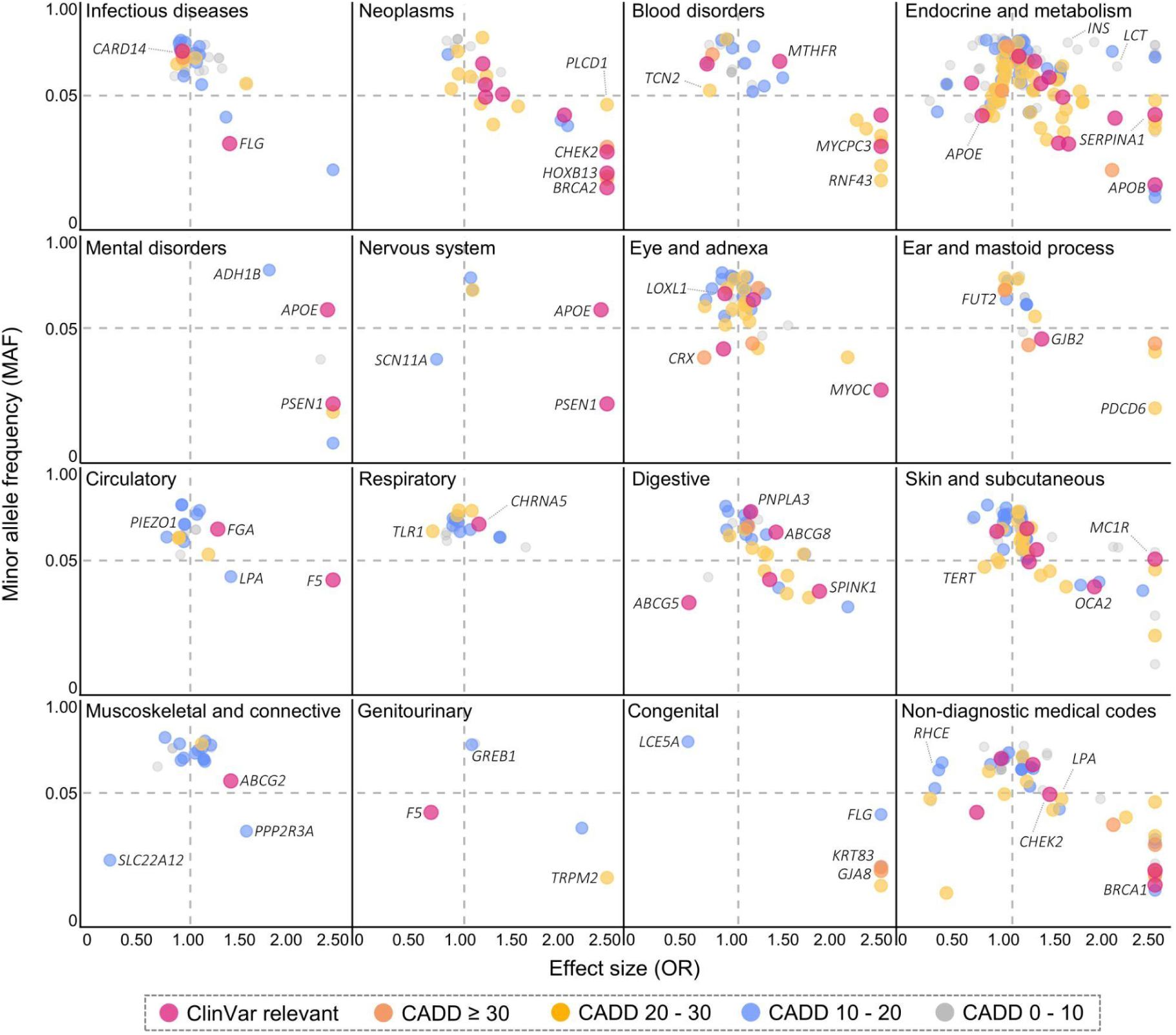
Scatterplots highlighting identified coding variants. Scatterplots show coding variants across ICD-10 code categories. The x-axis indicates the odds ratio, and the y-axis shows allele frequency on a log10 scale. Variants are coloured by prioritization class: pink, ClinVar-relevant; dark orange, CADD ≥ 30; light orange, CADD > 20; blue, CADD > 10; and gray, all remaining variants. Dashed reference lines mark an odds ratio of 1 and allele frequency of 0.05. Selected genes are annotated. Variant associations reaching genome-wide significance P < 5 × 10^-8^ are highlighted, with HLA locus variants having been excluded. OR - odds ratio.

A total of 417 variant-trait associations were inconclusive in fine-mapping, typically because local LD prevented separation of closely correlated candidates and PIP support remained distributed across multiple variants. This unresolved set included several established pathogenic loci, such as a rare stop-gained variant in *BRCA2*:p.Gln2858Ter (malignant neoplasm of breast, in females; ICD-10 C50; OR = 13.799; 95% CI = 6.948 - 27.407; *P* = 7.22 × 10^−14^), frameshift variants in *BRCA1*:p.Gln1756ProfsTer74 (malignant neoplasm of breast, in females; ICD-10 C50; OR = 14.175; 95% CI = 8.351 - 24.061; *P* = 9.10 × 10^−23^), *FLG*:p.Ser761CysfsTer36 (congenital ichthyosis; ICD-10 Q80; OR = 13.306; 95% CI = 8.806 - 20.107; *P* = 1.05 × 10⁻^34^), and the missense variant in *APOB*:p.Arg3527Gln (disorders of lipoprotein metabolism and other lipidaemias; ICD-10 E78; OR = 12.143; 95% CI = 8.438 - 17.474; *P* = 3.36 × 10^−41^). Such observations emphasize that lack of fine-mapping resolution does not preclude clinical relevance, and motivate prioritisation combining both statistical metrics and functional evidence.

### Population-scale mapping of HLA associations

Because of their extended linkage disequilibrium and strong effects, variants within the HLA locus were analysed separately from the main GWAS results. HLA alleles were imputed using SNP2HLA^32^ and tested across 1,369 major ICD-10 phenotypes. Among these, 55 major ICD-10 disease categories showed at least one Bonferroni-significant HLA association (*P* < 5 × 10^−8^), corresponding to 301 HLA-trait associations (**Supplementary Table 13**). For these categories, we performed follow-up analyses in their corresponding minor ICD-10 subcategories, identifying 87 additional phenotypes with significant HLA associations, yielding additional 443 HLA-trait associations at the same significance threshold (**Figure 5**).

**Figure 5.**
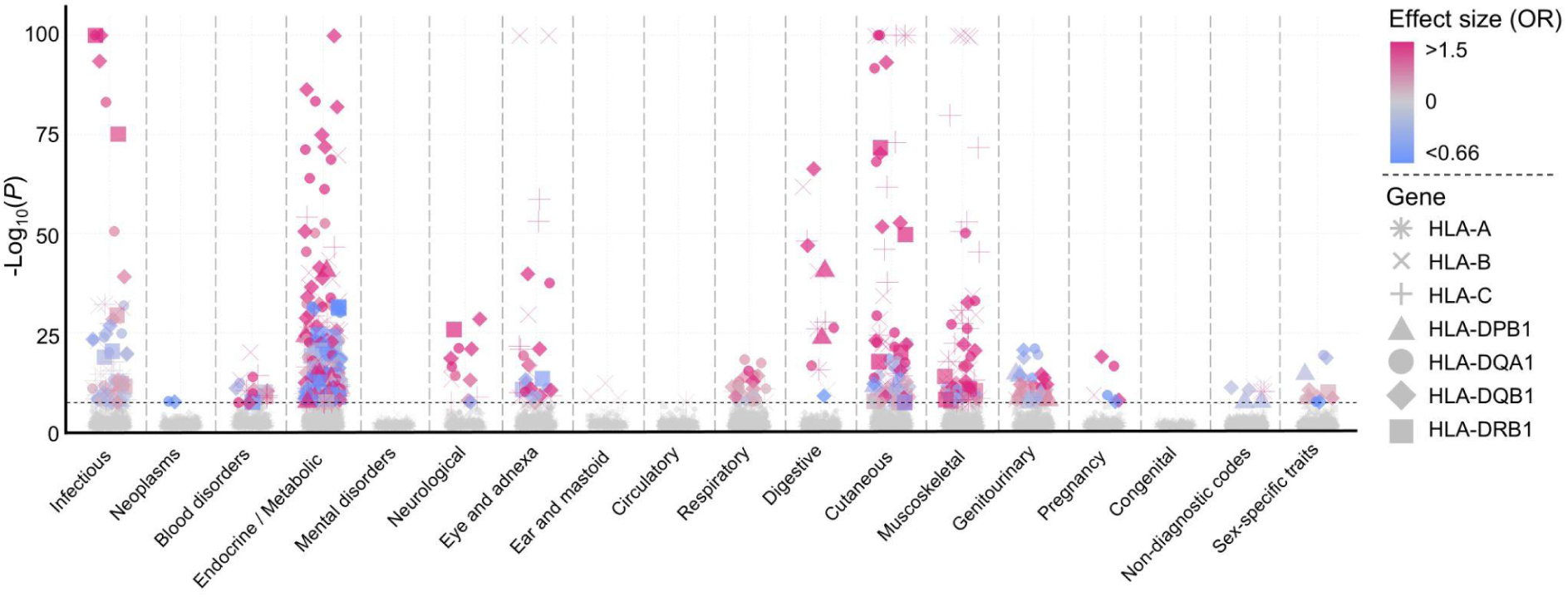
Scatterplot of HLA-phenotype association strength across ICD-10 trait groups. A total of 110 HLA protein structure-changing alleles were tested in a case control design consisting of ICD-10-based 1,612 traits. Each point represents one HLA allele tested against one ICD-10 phenotype, with the y-axis showing the association significance on a –log10(P) scale (capped at –log10(P) = 100). Points are grouped along the x-axis by broad ICD-10 trait chapters. Statistically significant associations (P < 5 × 10^−8^) are highlighted and colored by effect direction (blue for negative effects, pink for positive effects). Point shapes denote the underlying HLA genes. Sex-specific traits are shown as a separate category and include all phenotypes derived from male- or female-only analyses. The dashed horizontal line marks the significance threshold.

As expected, the strongest HLA signals were observed for infectious and immune mediated conditions. We observed well established HLA associations for classical immune mediated diseases, including thyroiditis (ICD-10 E06; 30 alleles), type 1 diabetes (ICD-10 E10; 18 alleles) and psoriasis (ICD-10 L40; 18 alleles). However, several outpatient-enriched phenotypes showed particularly robust HLA associations. These were especially prominent in inflammatory and autoimmune skin disorders, including seborrhoeic dermatitis (ICD-10 L21; 7 alleles), lichen planus (ICD-10 L43; 8 alleles), vitiligo (ICD-10 L80; 2 alleles), and psoriatic arthropathies (ICD-10 M07; 8 alleles). Strong HLA signals were also observed for common infectious phenotypes, such as viral warts (ICD-10 B07; 16 alleles), herpes zoster (ICD-10 B02; 8 alleles), and acute and chronic tonsillitis (ICD-10s J03 and J35.0, respectively; 5 alleles). In comparison to a similar recent HLA phenome-wide resource based on FinnGen project dataset^33^, our anlayses included six alleles observed only in our set (A*30:01, B*35:03, B*50:01, C*07:04, DPB1*15:01, DQA1*06:01), displaying biologically coherent associations among traits including herpesviral infections, thyroid disorders, acute and chronic tonsillitis, and psoriasis. At the phenotypic level, 45 ICD-10 traits were shared between datasets, while 97 were unique to EstBB. Notably, the trait rosacea (ICD-10 L71; 14,061 cases) displayed 10 HLA associations, of which 8 were putatively novel^34–37^ and included both protective and risk increasing effects (**Supplementary Figure 10**), illustrating how the exploratory resource presented here can complement and expand existing HLA association maps.

### Fungal infection host genetics and a *TNFSF15* splice signal

To illustrate how EstBB enables layered interpretation from a previously under-studied phenotype definition through fine-mapping and cross-biobank replication, we highlight pityriasis versicolor (ICD-10 B36.0). This skin disorder is caused by a common superficial fungal infection driven by the *Malassezia* genus^38^. In EstBB, it is primarily recorded in specialist care, with 22,674 of 26,181 diagnosis events (86.60%) originating from specialist encounters, supporting high diagnostic specificity. Our GWAS of 9,446 cases identified 31 independent loci and 14 significant HLA alleles (**Supplementary Figure 11, Supplementary Table 14**), implicating immune mediated susceptibility, with protein structure-altering variants present in established immune pathway genes including *HLA*, *SH2B3*, *PTPN22*, *IL13*, and *GSDMB*^39–41^. Notably, we also detected both common and rare associations near *CARD9*, *CARD11*, and *CARD14* genes, that have well established roles in skin inflammation^42^ and antifungal host defence^43^. Within the *TNFSF15* locus, fine-mapping and conditional analyses identified three independent signals, two driven by common variants and one led by a rare variant. The Estonian-specific rare variant rs1473488154 was one of the strongest putatively novel associations in the whole study by p-value (**Supplementary Figure 12**; OR = 7.633; 95% CI = 6.436 - 8.830; *P* = 1.45 × 10^−29^), supported by a high confidence in fine-mapping (PIP = 0.993). As a result of its high deleteriousness (CADD = 32) and predicted splice disruption (SpliceAI acceptor loss = 0.99; Pangolin splice loss = 0.83), this variant was prioritized for further analyses. The variant is located two basepairs upstream from exon 2 of *TNFSF15*, which is a gene encoding the tumor necrosis factor superfamily ligand TL1A. Exon 2 of *TNFSF15* encodes the cleavage site for proteolytic processing^44^, which regulates the balance between membrane bound inactive TL1A and soluble active form sTL1A (**Figure 6A**). The splicing predictions for rs1473488154 are consistent with exon 2 skipping, which is expected to produce an uncleavable and dysfunctional TL1A protein and could plausibly increase susceptibility or severity of fungal infections due to deficient immune reactions. Outside superficial mycoses, rs1473488154 showed no other statistically significant associations in PheWAS (**Supplementary Figure 13**), indicating a largely phenotype specific effect that may remain undetected in biobank datasets dominated by inpatient setting diagnoses.

**Figure 6:**
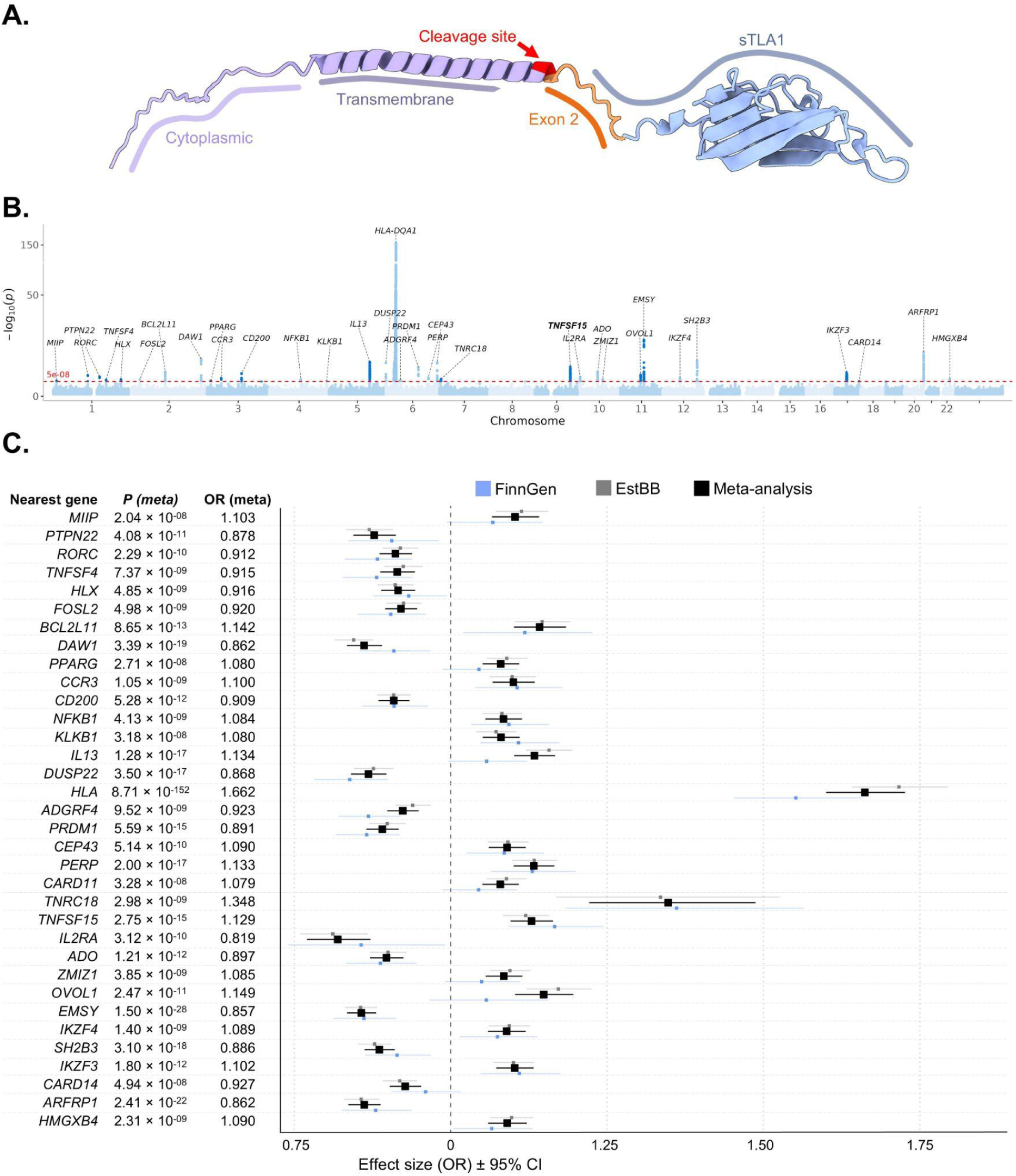
GWAS meta-analysis of pityriasis versicolor and TNFSF15 protein structure. **A** Schematic representation of the TNFSF15 protein structure. The protein contains a transmembrane domain and a globular extracellular domain. The region encoded by exon 2 is highlighted in orange. The cleavage site is indicated by a red arrow. **B** Manhattan plot of GWAS meta-analysis summary statistics for pityriasis versicolor. A total of 12,546,294 genetic variants (round dots) were tested in a case control design comprising 12,017 cases and 694,629 controls from EstBB and FinnGen biobanks (706,646 total). The x-axis shows genomic position in physical order, and the y-axis shows the strength of association as −log10(p-value). The red horizontal line indicates the genome-wide significance threshold (P = 5 × 10^-8^). Nearest genes to lead variants at associated loci are annotated. The TNFSF15 locus has been bolded. **C** Forest plot of lead variants across 34 loci associated with pityriasis versicolor in the EstBB and FinnGen meta-analysis. For each locus, the effect size of the genetic variant with the lowest p-value is shown. Effect estimates are displayed as odds ratios, with error bars representing 95% confidence intervals. Estimates from EstBB are shown in gray, FinnGen in blue, and the meta-analysis estimate in black.

Pityriasis versicolor displayed the highest SNP-based heritability among infectious diseases in this study (h^2^ = 0.132, *P* = 3.86 × 10^−12^), further supporting the substantial host genetic contribution to susceptibility for this common superficial fungal infection. GWAS analysis replication in FinnGen (2,571 cases) supported consistency of effects, with high cross-cohort genetic correlation (rg = 0.773, *P* = 1.70 × 10^−7^), and meta-analysis with a total of 706,646 participants increased the number of independent genome-wide significant loci to 34 (**Figure 6B-6C, Supplementary Table 15**). Gene set analyses supported the enrichment of immune mediated mechanisms, highlighting pathways involved in leukocyte and lymphocyte activation, cell activation, and T-cell differentiation (**Supplementary Table 16**). Overall, the example of pityriasis versicolor is consistent with prior evidence of shared genetic liability between dermatological and immune phenotypes^45,46^ and, to our knowledge, represents the first host-genetic GWAS report for this trait, while also highlighting an Estonian-enriched potential founder variant (**Supplementary Figure 14**). In conclusion, this study highlights the value of EstBB resources in uncovering previously understudied and underpowered disease phenotypes and mapping both common and rare genetic variant associations.

## Discussion

Conducting genome-wide association studies in well-phenotyped, population-specific biobanks enables the investigation of previously unexplored phenotypes and the identification of potential population-specific associations. Here, we report GWAS results for 5,491 ICD-10 phenotypes in 206,159 EstBB participants, highlighting how population biobanks with rich outpatient data can complement hospital-focused resources and enable genetic analyses of underrepresented clinical presentations. The publicly available summary statistics expand the landscape of clinically relevant associations and provide a foundation for downstream work on mechanisms, disease stratification, and risk prediction.

Our analysis demonstrates that the Estonian Biobank adds value to largely hospital-centred biobanks through its enriched primary care data, as observed in comparison with the UK Biobank, while yielding results comparable to those obtained from genetic correlation analyses with FinnGen. Most associations in our study replicate prior evidence as well - we identified 689 (98.43%) study-wide significant loci to be present at least in the GWAS Catalog, OpenTargets Genetics or OpenTargets Platform^47–49^, supporting the robustness of our approach. At the same time, EstBB enabled discovery in outpatient-rich traits that previously have seen limited genetic study. To illustrate the novelty enabled by such data, we highlight pityriasis versicolor for which 34 largely immune-related loci were identified, representing the first GWAS of this trait. Notably, the lead HLA signal, HLA-DQB1*05:01 (OR = 1.693; *P* = 1.0 × 10^−122^), has previously been implicated in sensitization to low-molecular-weight organic acid anhydrides^50^, raising the possibility that the *Malassezia* produced metabolite pityriaanhydride could contribute to heightened local immune reactivity in susceptible individuals. Pityriasis versicolor is a specialist-captured phenotype that revealed a high-quality genetic signal, including a splice-disrupting *TNFSF15* variant consistent with altered TL1A signalling in host defence. Given that pharmacological inhibition of related inflammatory pathways is associated with increased fungal susceptibility^51^, this finding suggests that population-enriched coding variants can act as natural experiments that model therapeutic modulation of immune pathways.

A substantial fraction of the variants identified here mapped to ICD-10 subcategories, which can function as clinically meaningful endophenotypes that refine broad diagnoses into more specific etiological entities. Previous studies in population-based collections have demonstrated similar results, illustrating the benefits of analysing homogeneous population samples^10^. Furthermore, the trait granularity provides a practical framework for re-examining comorbidity structure and for identifying shared and distinct genetic components across closely related clinical presentations.

Notwithstanding the relevance of our findings, several limitations merit consideration, especially with respect to phenotype definition and ascertainment. ICD-10–based phenotypes provide broad coverage but are inherently heterogeneous and shaped by clinical practice. Defining cases based on a single recorded event can increase statistical power, but it may also capture transient diagnoses and introduce misclassification, thereby inflating association signals. For example, the phenotype desensitisation to allergens (ICD-10 Z51.6) likely reflects pregnancy-related rhesus prophylaxis rather than allergy treatment, consistent with its strong female bias. As a result, phenotype-specific associations should be interpreted cautiously, as healthcare-system practices can generate spurious signals. Greater specificity could be achieved by requiring repeated diagnostic codes, incorporating prescription or procedure data, or mapping ICD-10 codes to harmonised groupings such as Phecodes^52^. For variant interpretation, fine-mapping improves locus resolution; however, some coding associations remain difficult to interpret due to linkage disequilibrium or low allele frequencies and will require functional follow-up.

Overall, our results show that outpatient-rich population biobanks can extend genetic discovery into understudied phenotypes, refine disease architecture through granular clinical endpoints, and reveal coding variants that motivate mechanistic and translational follow-up. By mapping genotype-phenotype relationships at scale, this work contributes to an expanded catalogue of human disease associations and a more detailed taxonomy of clinical phenotypes that can support future precision medicine efforts.

## Methods

### Phenotype definitions

The Estonian Biobank is a volunteer-based biobank comprising 212,948 participants (data freeze 2026v02). All participants provided broad informed consent, allowing for the regular linkage of their health data with the National Health Insurance Fund and other relevant databases to obtain ICD-10 code based events. Most electronic health records have been collected since 2004^15^, and analyses were carried out with the data freeze 2023v01. Data from the national health databases of the Estonian Health Insurance Fund formed the primary source, contributing 43,877,635 records (63.68% of the total data). Text-mined information from medical epicrises followed, with 19,137,015 records (27.77%), while hospital and specialised registry datasets provided 5,339,037 medical events (7.75%) (**Figure 1A**). Regarding treatment types, outpatient care accounted for the most recorded interactions, with 63,393,173 cases. Inpatient care within secondary care facilities was represented with 4,212,773 cases. Analyses were restricted to participants of European ancestry, as non-European participants comprised only 0.31% of the Estonian Biobank. The final study population included 206,159 participants with more than five distinct ICD-10 codes recorded in their electronic health records. Phenotypes were retained for GWAS if they had more than 100 recorded cases. For each ICD-10 phenotype, cases were defined as participants with at least one corresponding ICD-10 event, and all remaining participants without that diagnosis were used as controls. Sex-specific phenotypes were defined as traits for which more than 90% of cases occurred in one sex.

### Genotyping and imputation

All Estonian Biobank participants have been genotyped at the Core Genotyping Lab of the Institute of Genomics, University of Tartu, using Illumina Global Screening Arrays v1, v2 and v3. All the genetic variants were imputed using an Estonian-specific imputation panel; 295,810 have been directly genotyped^19^. Samples were genotyped and PLINK format files were created using Illumina GenomeStudio v2.0.4. Individuals were excluded from the analysis if their call rate was < 95%, if they were outliers of the absolute value of heterozygosity (> 3SD from the mean), or if sex defined based on heterozygosity of the X chromosome did not match sex in phenotype data^19^. Before imputation, variants were filtered by call rate < 95%, Hardy-Weinberg equilibrium *P* < *10^−4^* (autosomal variants only), and minor allele frequency < 1%. Genotyped variant positions were in build 37 and were lifted over to build 38 using Picard. Phasing was performed using the Beagle v5.4 software^53^. Imputation was performed with Beagle v5.4 software (beagle.22Jul22.46e.jar) and default settings. The dataset was split into batches of 5,000. A population-specific reference panel consisting of 2,695 whole-genome sequenced samples was utilised for imputation, and standard Beagle hg38 recombination maps were used. Based on principal component analysis, samples not of European ancestry were removed. Duplicate and monozygous twin detection was performed with KING 2.2.7^54^, and one sample from each pair of duplicates was excluded.

### Genome-wide association studies

Association analysis in Estonian Biobank was carried out for all variants with an INFO score ≥ 0.4 for common variants and INFO score ≥ 0.8 for low-frequency variants (MAF < 0.05), using the additive model as implemented in REGENIE v3.2.1 with standard binary trait settings ^55^. The analysis was performed in two steps: first, it fits a null model using leave-one-chromosome-out (LOCO) cross-validation, and the second step performs association testing using Firth’s logistic regression for rare variant analysis to handle potential biases^56^. Logistic regression was carried out with adjustment for current age, age², sex, and 10 PCs as covariates, analysing only variants with a minimum minor allele count of 2. Before the GWAS, all the phenotypes were divided into non-sex-specific, male, and female sex-specific groups, which were analysed separately. Manhattan plots were generated using the topr R package^57^. After analyses, quality control was conducted to ensure that all the GWASs had the same number of genetic variants and chromosomes. For downstream PheWAS analyses, we extracted key variant-level results from the GWAS summary statistics, including allele frequency, effect size estimates and -log10(*P*).

### SNP-based heritability estimation

SNP-based heritability was estimated for each phenotype using LD Score Regression (LDSC v1.0.1) with European LD scores derived from the 1000 Genomes reference panel and restricting analyses to HapMap3 variants^58,59^. For binary traits, we calculated both observed-scale and liability-scale heritability, with liability-scale estimates adjusted for sample prevalence. Effective sample size was calculated as N_eff_ = 4/((1/N_cases_)+(1/N_controls_)) and used to assess the relationship between heritability estimates and case-control imbalance, following principles applied in previous UK Biobank analyses. Heritability *P* values were corrected using multiple testing (Bonferroni-corrected *P* < 9.11 × 10^-6^). Data was then categorised based on the Neff of less than 4,500, between 4,500 and 20,000, between 20,000 and 40,000, and 40,000 or higher. Corresponding to these conditions, h^2^ confidence labels “None,” “Low,” “Medium,” and “High” are assigned. If a row in the dataset does not meet the specified conditions, it is labelled “Unknown.” This classification was adapted from UKBB analysis from Neale’s lab, unpublished.

### Genetic correlation analysis

Genetic correlations were estimated using LD Score Regression (LDSC v1.0.1) for ICD-10 phenotypes with more than 5,000 cases and at least one genome-wide significant association, in total 684 non-sex-specific phenotypes. To maximise clinical resolution while avoiding duplicate representation of subcategory diagnoses, we analysed 4-digit ICD-10 codes where available and retained 3-digit codes only when no corresponding 4-digit subcategories were present. LDSC analyses were performed using European LD scores from the 1000 Genomes reference panel and restricting variants to the HapMap3 set^58,59^. Overall, 483 traits displayed significant genetic correlation with at least one other phenotype. We observed 9,309 Bonferroni-significant genetic correlations (P < 7.31 × 10^-5^), of which 2,870 occurred between phenotype pairs with limited co-occurrence at the record level (phenotypic odds ratio < 1.5), suggesting shared inherited liability beyond simple diagnostic overlap.

For cross-biobank genetic correlation analyses with FinnGen, we used publicly available summary statistics from the FinnGen r12 release and selected traits with clinically comparable endpoints between EstBB ICD-10 phenotypes and FinnGen curated traits^33^. Traits were required to be diagnosis based, to have broadly comparable clinical definitions across cohorts, to include more than 5,000 cases where possible, and to show adequate GWAS signal, defined here as at least two independent genome-wide significant loci. After applying these criteria, 100 traits from distinct ICD-10 categories were retained for cross-biobank LDSC analyses.

### Phenotypic correlation analysis

Phenotypic overlap between individual ICD-10 traits was assessed using pairwise logistic regression in the Estonian Biobank. Each ICD-10 phenotype was analysed as a binary outcome, with a second phenotype included as the predictor in the same model. All analyses were adjusted for sex, age, age squared, year of birth, and the first five genetic principal components. To maintain consistency with the genetic correlation analyses, we restricted this analysis to phenotypes with more than 5,000 cases, matching the lower case-count threshold applied in LDSC genetic correlation analyses. Phenotypic overlap odds ratios of at least 1.5 were considered to indicate strong positive overlap.

### Lead variant identification and loci definition

A window size of ±500 kb was used to determine lead SNPs, ensuring that the identified lead SNPs were independent. For each genome-wide significant region (P = 5 × 10^-8^), the variant with the highest absolute Z-score has been designated as the lead variant, with MAF > 0.001. If the loci defined during lead-variant estimation overlapped, they were merged into larger loci, with a maximum locus size of 6 Mb. The HLA region was defined on chromosome 6, from 25,000,000 to 35,000,000 bp (hg38) and excluded from the analyses. If the locus overlapped with the HLA region, it was clipped to the HLA region’s border. Study-wide significance threshold was defined by applying the Bonferroni correction (5 × 10^-8^ / 5,000, where 5,000 is the approximate number of traits in the analysis).

### Fine-mapping

For each identified locus, fine mapping was performed using SuSie fine-mapping with the SuSieR v0.14.2 R package^60^. In-sample LD matrices have been calculated with LDStore v2.0^61^ as part of the pipeline and used for the analysis. LD matrices, together with GWAS summary statistics, were then used to conduct SuSie fine-mapping with the following settings: maximum number of causal variants in a locus L = 10, min_cs_corr = 0.5, and coverage = 0.99. For further analyses, the results for 99% probability credible sets have been used.

### Phenotype and locus comparisons with UKBB

To compare EstBB with UKBB, we first assessed ICD-10 phenotype availability and case counts using UKBB Data-Field 41202 (https://biobank.ndph.ox.ac.uk/showcase/field.cgi?id=41202&nl=1), which contains main ICD-10 diagnosis counts linked to hospital records. EstBB ICD-10 traits were matched to the corresponding UKBB ICD-10 codes, and case and control counts were compared across all codes observed in either resource. To compare the EstBB GWAS results with UKBB^62^ we used the summary statistics downloaded as flat files from pan-UKBB (https://pan.ukbb.broadinstitute.org). We extracted the associations for GWAS conducted in Europeans while confining ourselves to variants of high confidence in Europeans. After overlap with ICD-10 codes studies in our EstBB GWAS, this yielded 771 broad ICD-10 codes available for comparison. European-specific case counts were extracted for the Pan-UKBB phenotype manifest file. To count the number of genome-wide associated loci (*P* < 5 × 10^-8^), we applied the same locus definition as in our EstBB GWAS.

### Known association lookup

To distinguish the loci not previously described publicly, we identified putatively novel loci by overlapping the credible set variants from our analysis with previously known genetic associations from the GWAS Catalog (version v1.0.2, *P* < 1 × 10^-5^)^48^, OpenTargets Genetics (*P* < 5 × 10^-3^), and the OpenTargets Platform (*P* < 1 × 10^-4^)^49^. After the credible set variants were identified, they were overlapped with the variants in the GWAS Catalog based on the combination of the chromosome and position of each genetic variant. All variants were overlapped with OpenTargets Genetics and OpenTargets Platform based on the combination of the chromosome, position, reference and alternative alleles. The ones present in the GWAS Catalog dataset were considered “Known,” and those unique to our dataset were labelled “Novel.” If at least one SNP per credible set was considered to be “Known”, the locus was considered to be “Known” as well. This approach was adapted from Verma et al. (2024)^47^. For the loci that did not produce a credible set during fine-mapping, we retained the lead variant defined as the variant with the most significant absolute *z*-score (*P* < 5 × 10⁻⁸). These lead variants were subsequently assessed against the same reference databases, and two were not reported to have genome-wide significant associations and were therefore considered putatively novel.

### HLA imputation and association testing

HLA imputation of the Estonian Biobank genotype data was performed at the Broad Institute using the SNP2HLA tool^32^. The imputation was done for genotype data generated on the Global Screening Arrays v1, v2 and v3. We performed separate additive logistic regression analysis with the imputed HLA alleles using REGENIE v4.0 with standard binary trait settings^55^. Logistic regression was carried out with adjustment for current age, age², year of birth, sex and 5 PCs as covariates, using data ICD-10 based phenotypic data from EstBB 2025v01 freeze. Analyses were performed for all ICD-10 diagnoses meeting 100 cases threshold, and association significance was evaluated using a strict GWAS-wide threshold corresponding to *P* < 5 × 10^-8^. For ICD-10 categories, we first tested broad category-level phenotypes (such as E10 - “Type 1 diabetes mellitus”). For categories with Bonferroni-significant HLA signals, we subsequently evaluated corresponding more granular ICD-10 subcategories (such as E10.0 - “Type 1 diabetes mellitus with coma”). To limit redundancy and focus on functionally interpretable signals, we report results for classical HLA alleles at the two-field resolution (for example A*01:01). In the text we report only associations passing the chosen significance threshold and with imputation confidence INFO ≥ 0.9, while full results for all tested alleles and phenotypes, including non-significant associations and synonymous or serogroup-level mappings, are provided in the source data.

To compare the EstBB HLA association results against an external resource, we compared the dataset against a recently published FinnGen HLA phenome-wide analysis^33^. We matched HLA allele identifiers by allele names at the same locus and two-field resolution, and quantified overlap in the set of alleles available for association testing. Phenotype overlap was assessed by matching ICD-10 codes to the definitions used in the external resource, and traits were classified as shared or cohort-specific based on their inclusion in the respective analysed trait sets. For shared alleles and phenotypes, we compared the direction and strength of association at the predefined significance threshold.

### Coding variant annotation and analysis

Genome-wide significant variants from all GWAS analyses were cross-referenced with exonic variants to identify coding variants associated with phenotypic traits. For this, we used a genome-wide significance threshold of *P* < 5 × 10⁻^8^. Exonic region limits were obtained from gnomAD v4.1.0^63^, which includes 730,947 exomes and 76,215 whole genomes mapped to the GRCh38 reference genome. To characterise the functional consequences of coding variants, we used the Variant Effect Predictor^64^, integrating annotations from ClinVar and CADD^65^ to assess potential pathogenicity. ClinVar relevant variants were defined as coding variants annotated in ClinVar as pathogenic, conflicting classifications of pathogenicity, drug response, or risk factor. Protein structure- or function-altering variants were defined as those annotated with at least one of the following consequence terms: missense, stop_gained, frameshift and inframe_deletion.

### Pityriasis versicolor analysis

For the pityriasis versicolor case study, we re-ran the GWAS using the latest available EstBB phenotype freeze (2026v01) to maximise case ascertainment and statistical power. Cases were defined as participants with at least one recorded ICD-10 diagnosis of B36.0, yielding 9,446 cases. Controls were defined as genotyped EstBB participants without any recorded B36.0 diagnosis in the linked health records, resulting in 196,675 controls and 206,121 individuals in total. Association testing was performed with REGENIE v4.0 under a binary trait model, adjusting for sex, age, age^2^, year of birth, and the first 10 genetic principal components. Replication was carried out using FinnGen summary statistics for pityriasis versicolor, where cases were defined by ICD-10 code B36.0 (2,571 cases) and all participants without the diagnosis were similarly used as controls (497,954 controls). Cross-biobank meta-analysis of EstBB and FinnGen results was performed using GWAMA^66^, combining in total 12,017 pityriasis versicolor cases. Because EstBB and FinnGen represent genetically related Northern European populations, we applied relatively lenient allele frequency filters of MAF > 0.001 and MAF < 0.99 to retain shared low-frequency variation and improve power for rare variant discovery. Independent loci were defined from the meta-analysis results using the same locus definition and clumping strategy as in the main GWAS analyses. To summarise biological pathways implicated by the pityriasis versicolor loci, we performed gene set enrichment analysis using g:Profiler^67^, where input genes were defined by mapping each lead genetic variant to its nearest gene. For the TNFSF15 protein structure, we used the precomputed AlphaFold model AF-O95150-F1-v6^68,69^ and modified the structure for visual clarity. Structural models were visualized in UCSF ChimeraX v1.10.1^70^.

## Ethics statement

The activities of the EstBB are regulated by the Human Genes Research Act, which was adopted in 2000 specifically for the operations of EstBB. Individual level data analysis in EstBB was carried out under ethical approval 1.1-12/624 from the Estonian Committee on Bioethics and Human Research (Estonian Ministry of Social Affairs), using data according to release application 3-10/GI/31689 from the Estonian Biobank.

Study subjects in FinnGen provided informed consent for biobank research, based on the Finnish Biobank Act. Alternatively, separate research cohorts, collected prior the Finnish Biobank Act came into effect (in September 2013) and start of FinnGen (August 2017), were collected based on study-specific consents and later transferred to the Finnish biobanks after approval by Fimea (Finnish Medicines Agency), the National Supervisory Authority for Welfare and Health. Recruitment protocols followed the biobank protocols approved by Fimea. The Coordinating Ethics Committee of the Hospital District of Helsinki and Uusimaa (HUS) statement number for the FinnGen study is Nr HUS/990/2017.

The FinnGen study is approved by Finnish Institute for Health and Welfare (permit numbers: THL/2031/6.02.00/2017, THL/1101/5.05.00/2017, THL/341/6.02.00/2018, THL/2222/6.02.00/2018, THL/283/6.02.00/2019, THL/1721/5.05.00/2019 and THL/1524/5.05.00/2020), Digital and population data service agency (permit numbers: VRK43431/2017-3, VRK/6909/2018-3, VRK/4415/2019-3), the Social Insurance Institution (permit numbers: KELA 58/522/2017, KELA 131/522/2018, KELA 70/522/2019, KELA 98/522/2019, KELA 134/522/2019, KELA 138/522/2019, KELA 2/522/2020, KELA 16/522/2020), Findata permit numbers THL/2364/14.02/2020, THL/4055/14.06.00/2020, THL/3433/14.06.00/2020, THL/4432/14.06/2020, THL/5189/14.06/2020, THL/5894/14.06.00/2020, THL/6619/14.06.00/2020, THL/209/14.06.00/2021, THL/688/14.06.00/2021, THL/1284/14.06.00/2021, THL/1965/14.06.00/2021, THL/5546/14.02.00/2020, THL/2658/14.06.00/2021, THL/4235/14.06.00/2021, Statistics Finland (permit numbers: TK-53-1041-17 and TK/143/07.03.00/2020 (earlier TK-53-90-20) TK/1735/07.03.00/2021, TK/3112/07.03.00/2021) and Finnish Registry for Kidney Diseases permission/extract from the meeting minutes on 4^th^ July 2019.

The Biobank Access Decisions for FinnGen samples and data utilized in FinnGen Data Freeze 12 include: THL Biobank BB2017_55, BB2017_111, BB2018_19, BB_2018_34, BB_2018_67, BB2018_71, BB2019_7, BB2019_8, BB2019_26, BB2020_1, BB2021_65, Finnish Red Cross Blood Service Biobank 7.12.2017, Helsinki Biobank HUS/359/2017, HUS/248/2020, HUS/430/2021 §28, §29, HUS/150/2022 §12, §13, §14, §15, §16, §17, §18, §23, §58, §59, HUS/128/2023 §18, Auria Biobank AB17-5154 and amendment #1 (August 17 2020) and amendments BB_2021-0140, BB_2021-0156 (August 26 2021, Feb 2 2022), BB_2021-0169, BB_2021-0179, BB_2021-0161, AB20-5926 and amendment #1 (April 23 2020) and it’s modifications (Sep 22 2021), BB_2022-0262, BB_2022-0256, Biobank Borealis of Northern Finland_2017_1013, 2021_5010, 2021_5010 Amendment, 2021_5018, 2021_5018 Amendment, 2021_5015, 2021_5015 Amendment, 2021_5015 Amendment_2, 2021_5023, 2021_5023 Amendment, 2021_5023 Amendment_2, 2021_5017, 2021_5017 Amendment, 2022_6001, 2022_6001 Amendment, 2022_6006 Amendment, 2022_6006 Amendment, 2022_6006 Amendment_2, BB22-0067, 2022_0262, 2022_0262 Amendment, Biobank of Eastern Finland 1186/2018 and amendment 22§/2020, 53§/2021, 13§/2022, 14§/2022, 15§/2022, 27§/2022, 28§/2022, 29§/2022, 33§/2022, 35§/2022, 36§/2022, 37§/2022, 39§/2022, 7§/2023, 32§/2023, 33§/2023, 34§/2023, 35§/2023, 36§/2023, 37§/2023, 38§/2023, 39§/2023, 40§/2023, 41§/2023, Finnish Clinical Biobank Tampere MH0004 and amendments (21.02.2020 & 06.10.2020), BB2021-0140 8§/2021, 9§/2021, §9/2022, §10/2022, §12/2022, 13§/2022, §20/2022, §21/2022, §22/2022, §23/2022, 28§/2022, 29§/2022, 30§/2022, 31§/2022, 32§/2022, 38§/2022, 40§/2022, 42§/2022, 1§/2023, Central Finland Biobank 1-2017, BB_2021-0161, BB_2021-0169, BB_2021-0179, BB_2021-0170, BB_2022-0256, BB_2022-0262, BB22-0067, Decision allowing to continue data processing until 31^st^ Aug 2024 for projects: BB_2021-0179, BB22-0067,BB_2022-0262, BB_2021-0170, BB_2021-0164, BB_2021-0161, and BB_2021-0169, and Terveystalo Biobank STB 2018001 and amendment 25^th^ Aug 2020, Finnish Hematological Registry and Clinical Biobank decision 18^th^ June 2021, Arctic biobank P0844: ARC_2021_1001.

## Funding statement

This work was funded by the European Union through Horizon 2020 and Horizon Europe research and innovation program under grants no. 874627 (EXPANSE) (JK); 894987 (GENOMEPEP) (EA); 101153901 (CHRONOPIA) (TP); 101096888 (DISCERN) (JK); 101137201 (CLARITY) (EA, KB, LT); 101057721 (PROPHET) (AR); 101128023 (JAPreventNCD) (AR); 101080009 (CAN.HEAL) (AR); 101137278 (CVDLINK) (UV); 101060011 (TeamPerMed) (Estonian Biobank Research Team and Health Informatics Research Team); University of Tartu Development Fund bridging grant PLTGIARENG24 (EA); Estonian Research Council Grants PRG555 (Evaluation of genetic variants potentially linked to actionable health risks) (AR); PRG1291 (Systematic phenome-wide search for genetic modulators in health and disease) (AA, EA, KB, JK, UV, PP), PSG1230 (Utilizing large-scale blood gene expression genetics data for causal gene prioritization and predictive modeling of traits and diseases) (UV), PRG1844 (Discovery and analysis of clinical pathways in health Data) (Health Informatics Research Team), Roadmap II project number TT17 (EstBB Research Team), and co-funded by the Ministry of Education and Research grant TEM-TA72 (Health Informatics Research Team).

The FinnGen project is funded by two grants from Business Finland (HUS 4685/31/2016 and UH 4386/31/2016) and the following industry partners: AbbVie Inc., AstraZeneca UK Ltd, Biogen MA Inc., Bristol Myers Squibb Inc. (and Celgene Corporation & Celgene International II Sàrl), Genentech Inc., Merck Sharp & Dohme LCC, Pfizer Inc., GlaxoSmithKline Intellectual Property Development Ltd., Sanofi US Services Inc., Maze Therapeutics Inc., Johnson&Johnson Innovative Medicine Inc., Novartis AG, Boehringer Ingelheim International GmbH and Bayer AG. Following biobanks are acknowledged for delivering biobank samples to FinnGen: Auria Biobank (https://www.auria.fi/biopankki), THL Biobank (https://www.thl.fi/biobank), Helsinki Biobank (https://www.helsinginbiopankki.fi), Biobank Borealis of Northern Finland (https://www.ppshp.fi/Tutkimus-ja-opetus/Biopankki/Pages/Biobank-Borealis-briefly-in-English.aspx), Finnish Clinical Biobank Tampere (https://www.tays.fi/en-US/Research_and_development/Finnish_Clinical_Biobank_Tampere), Biobank of Eastern Finland (https://www.ita-suomenbiopankki.fi/en), Central Finland Biobank (https://www.ksshp.fi/fi-FI/Potilaalle/Biopankki), Finnish Red Cross Blood Service Biobank (https://www.veripalvelu.fi/verenluovutus/biopankkitoiminta), Terveystalo Biobank (https://www.terveystalo.com/fi/Yritystietoa/Terveystalo-Biopankki/Biopankki/) and Arctic Biobank (https://www.oulu.fi/en/university/faculties-and-units/faculty-medicine/northern-finland-birth-cohorts-and-arctic-biobank). All Finnish Biobanks are members of BBMRI.fi infrastructure (https://www.bbmri-eric.eu/national-nodes/finland/). Finnish Biobank Cooperative -FINBB (https://finbb.fi/) is the coordinator of BBMRI-ERIC operations in Finland. The Finnish biobank data can be accessed through the Fingenious® services (https://site.fingenious.fi/en/) managed by FINBB.

## Acknowledgements

We want to acknowledge the participants of the Estonian Biobank for their contributions. The Estonian Genome Centre analyses were partially carried out in the High Performance Computing Center, University of Tartu. This work was written at writing retreats and writing days organised by the Institute of Genomics, University of Tartu.

The Estonian Biobank Research Team was responsible for data collection, genotyping, quality control and imputation and consisted of Andres Metspalu, Mait Metspalu, Lili Milani, Reedik Mägi, Mari Nelis and Georgi Hudjashov.

The Health Informatics Research Team was responsible for data harmonization, mapping to OMOP CDM, fact extraction of laboratory measurements and consisted of Raivo Kolde, Sven Laur, Sulev Reisberg and Jaak Vilo.

We also thank Andres Veidenberg, Abdelrahman Elnahas and Kadri Reis for their contribution in this work. We also want to acknowledge the participants and investigators of the FinnGen study.

## Data availability

GWAS summary statistics will be made publicly available upon publication in a peer-reviewed journal. Pseudonymised data and/or biological samples can be accessed for research and development purposes in accordance with the Estonian Human Genome Research Act (https://www.riigiteataja.ee/en/eli/ee/531102013003/consolide/current). To access the raw data, the research proposal must be approved by the Scientific Advisory Committee of the Estonian Biobank as well as by the Research Ethics Committee (https://etag.ee/en/research-culture/research-integrity/research-ethics-committee). For more details on data access and relevant documents, please see https://genomics.ut.ee/en/content/estonian-biobank#dataaccess.

## Code availability

All software programs used for the analyses described in this paper are freely available online: REGENIE v3.2.1 (https://github.com/rgcgithub/regenie); UCSC Genome Browser for variant analysis (http://genome.ucsc.edu/); Statistical analysis were carried out using R version 4.2.1; LDSC v1.0.1 (https://github.com/bulik/ldsc); pipelines were built using Nextflow v22.04.3; visualization of the protein structures and docking results using ChimeraX v1.10.1 (https://www.rbvi.ucsf.edu/chimerax/); LDSTORE for ld matrices calculation (https://stephenslab.github.io/susieR/articles/finemapping_summary_statistics.html); fine-mapping with SuSieR v0.14.2 (https://stephenslab.github.io/susieR/articles/finemapping_summary_statistics.html); For FinnGen, code to perform GWAS analyses is available at the FinnGen GitHub (https://github.com/FINNGEN/). Individual plots were created using R v4.2.1, including the R packages ggplot2, geojsonio, dplyr, and topr. Map in **Supplementary Figure 14** was generated using publicly available GeoJSON data from the Natural Earth dataset. The final figures were edited with Microsoft Powerpoint.

## Author contributions

A.A., E.A., J.K., U.V., T.E and P.P. designed the study.

The EstBB Research Team, Health Informatics Research Team and FinnGen project consortium collected and provided the genotype and phenotype data.

A.A., E.A., K.B., K.J., M.J., H.H., S.S., L.T., U.V., S-O.S., J.K. performed the analyses.

A.R. and T.P. assisted with clinical phenotype definitions.

U.V. and A.A. developed fine-mapping and variant identification pipelines.

H.M.O., U.V., T.E and P.P. supervised the study.

A.A., E.A., wrote the first draft of the manuscript. All authors critically reviewed the manuscript.

## Competing Interest Statement

The authors declare that they have no competing interests.

---

**Supplementary Figure 1:**
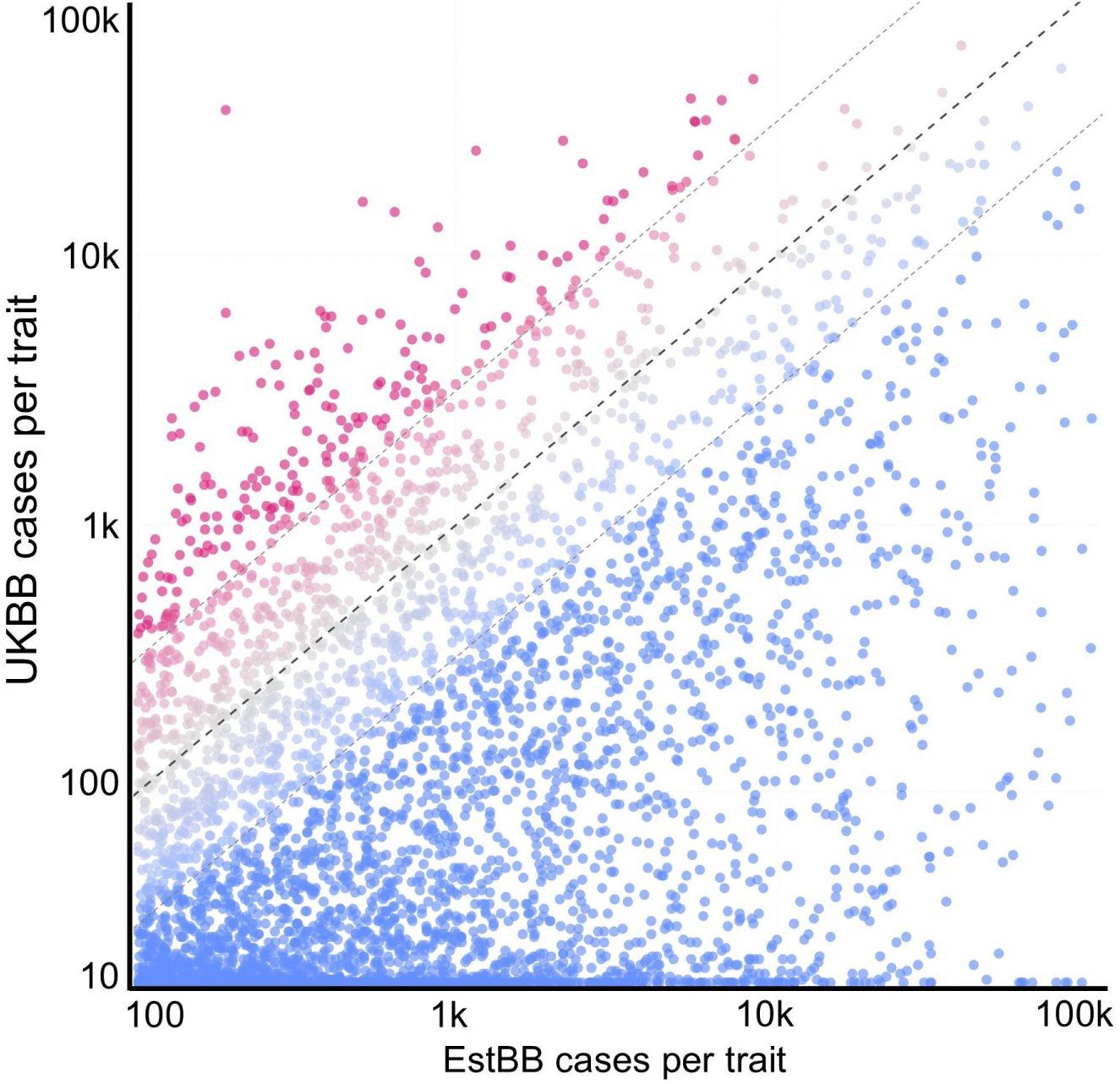
Comparison of Estonian Biobank and UK Biobank case counts across overlapping ICD-10 traits. Each point represents one ICD-10 phenotype shared between EstBB and UKBB, with 3,909 overlapping traits included. The x-axis shows EstBB case counts and the y-axis shows UKBB case counts, both on a log-transformed scale. The dashed diagonal indicates equal case numbers in the two biobanks. Point color reflects the ratio of EstBB to UKBB case counts, ranging from pink for UKBB-enriched traits, through gray for traits with similar case counts, to blue for EstBB-enriched traits. Only phenotypes with at least 100 cases in EstBB were included.

**Supplementary Figure 2:**
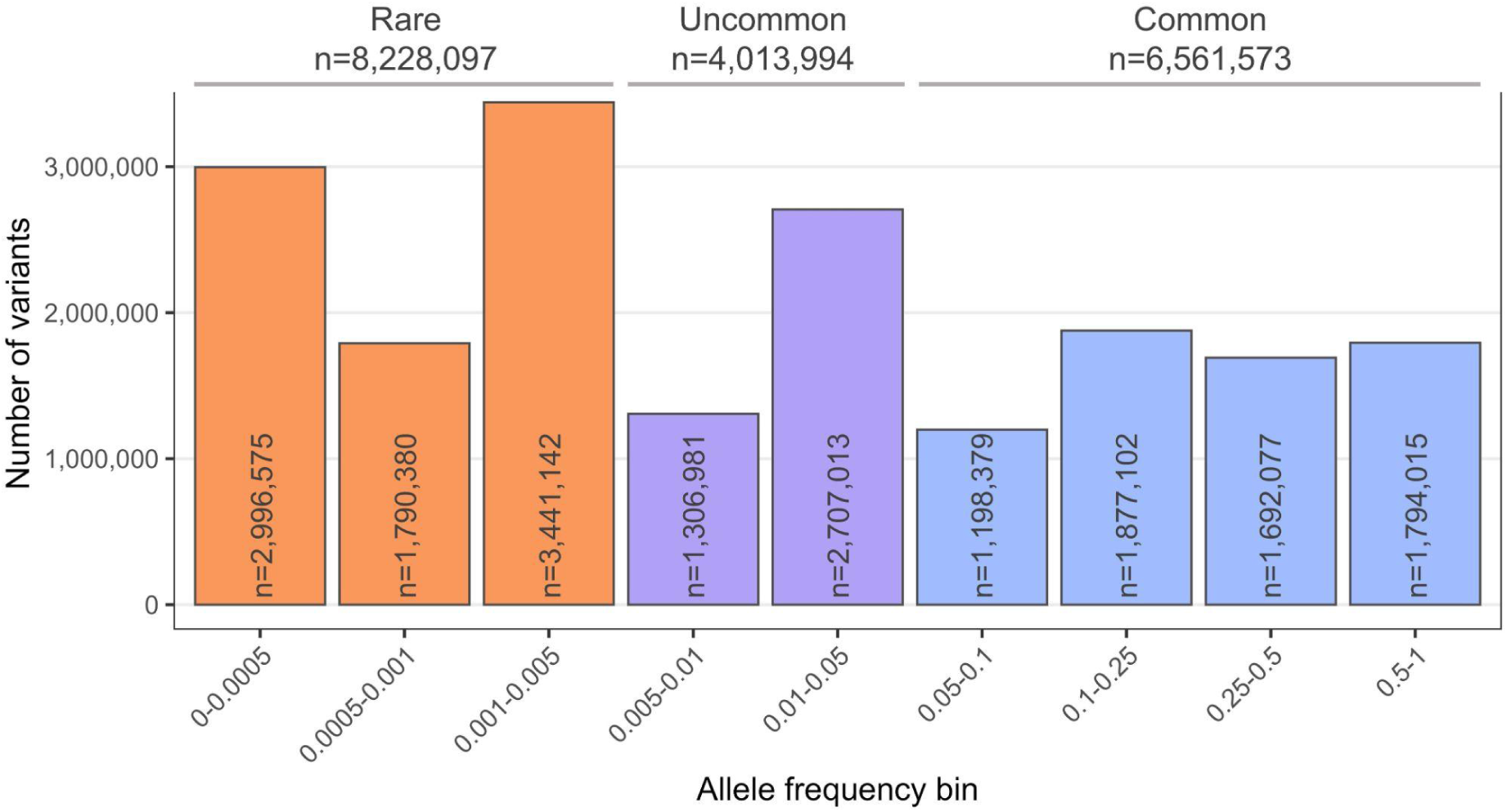
Distribution of variants included in the study across allele frequency bins. Association analysis in Estonian Biobank was carried out for all variants with an INFO score ≥ 0.4 for common variants and INFO score ≥ 0.8 for low-frequency variants (MAF < 0.05). Bars in the histogram show the number of variants within each frequency bin, with colours indicating rare, uncommon, and common frequency classes. Numbers inside the bars indicate the variant count in each bin. In total, 1,561,235 insertion-deletion variants and 17,242,429 single nucleotide variants were included, corresponding to 18,803,664 variants overall.

**Supplementary Figure 3:**
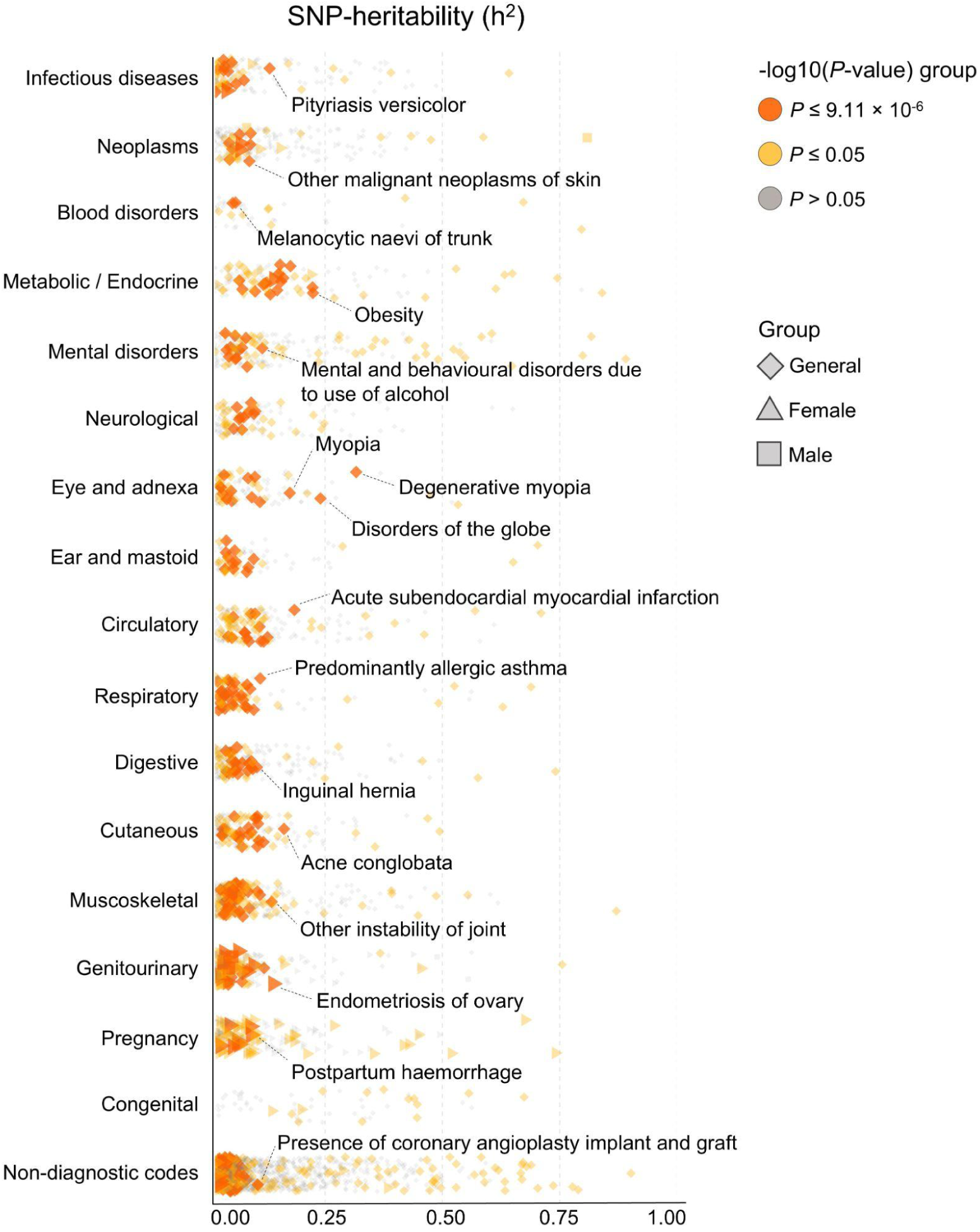
SNP heritability estimates across 5,491 GWAS traits. Scatter plot of SNP heritability estimates (h^2^) from LD score regression across 5,491 GWAS traits in EstBB, grouped by ICD-10 disease category. Each point represents one trait and is plotted by estimated h^2^ on the x-axis and disease category on the y-axis. Point colour indicates the significance group of the heritability estimate, corresponding to Bonferroni-significant (orange; P < 9.11 × 10^−6^), nominally significant (yellow; P ≤ 0.05), or non-significant results (gray; P > 0.05). Point shape indicates whether the trait was analysed in both sexes (rhombus), females only (triangle), or males only (square). Selected traits with notable heritability estimates are annotated.

**Supplementary Figure 4:**
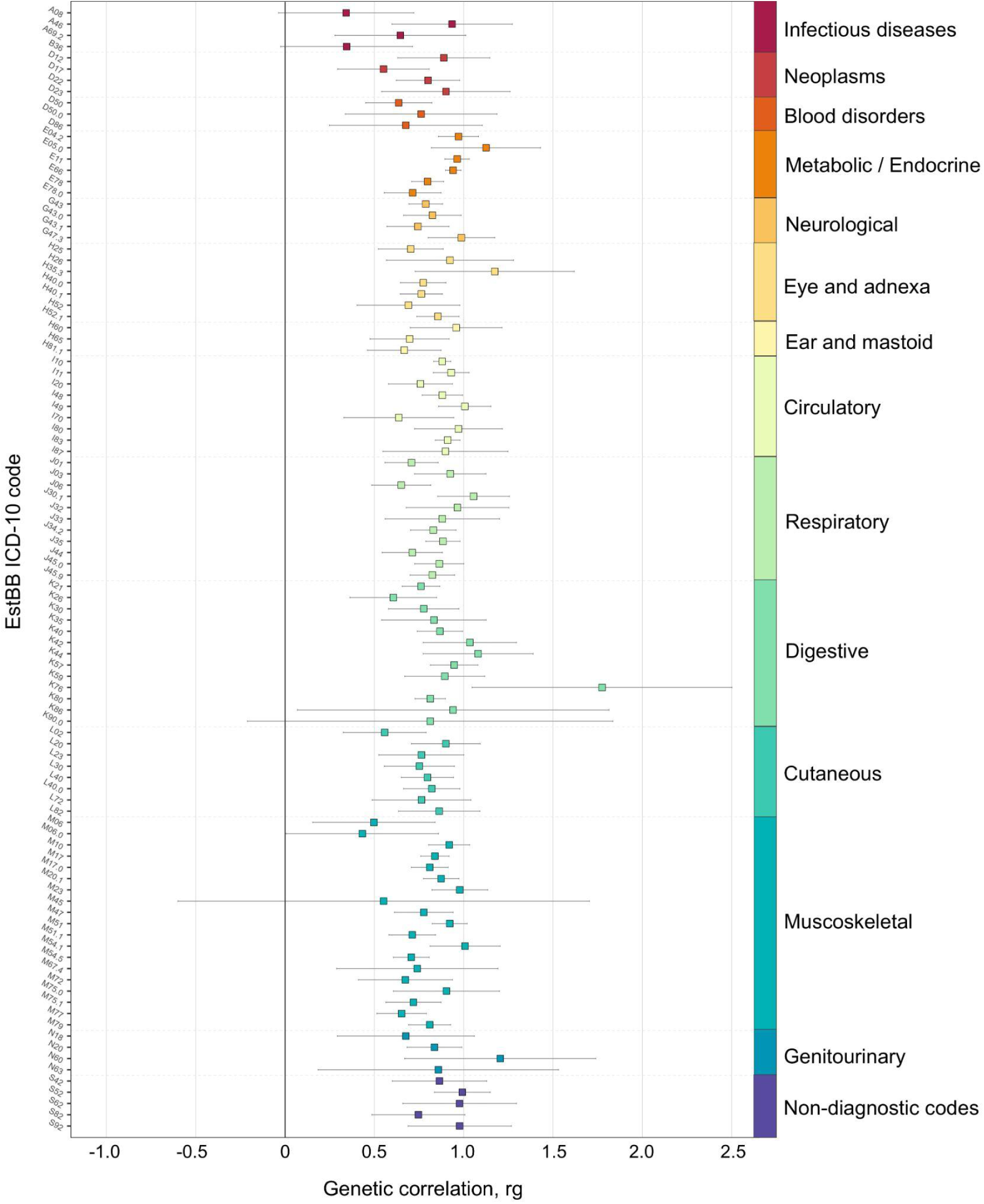
Comparison of genetic correlation estimates across 100 traits in Estonian Biobank and FinnGen. Genetic correlation estimates between Estonian Biobank and FinnGen for 100 ICD-10 traits with comparable phenotype definitions in both biobanks. Each square represents the LD score regression estimate for one trait, plotted by EstBB ICD-10 code on the x-axis and genetic correlation (rg) on the y-axis. Colours indicate ICD-10 disease categories, and error bars show 95% confidence intervals.

**Supplementary Figure 5:**
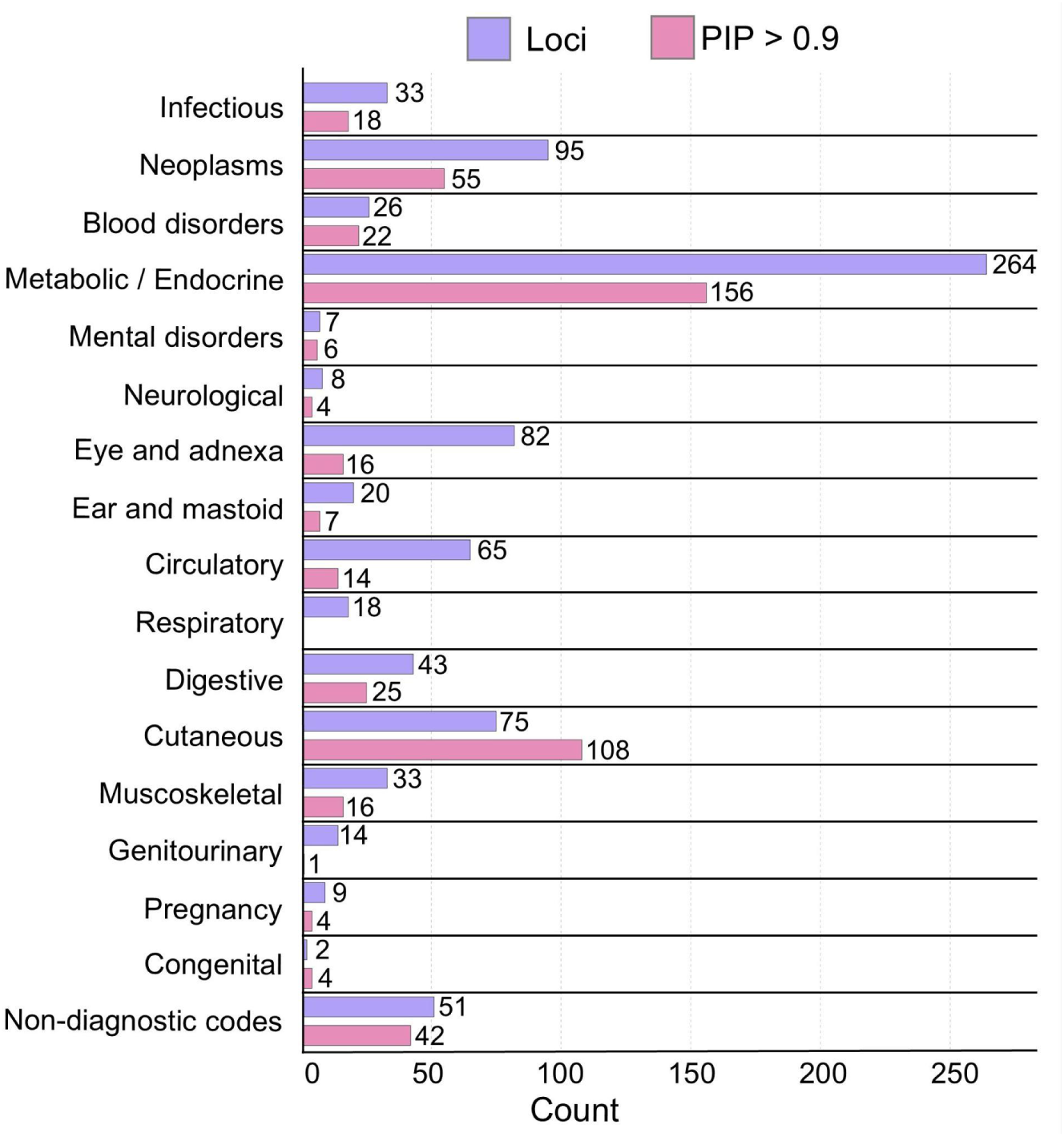
Counts of fine-mapped signals and associations across ICD-10 disease categories. The panel summarizes the number of significant locus-trait pairs per category reaching P < 1 × 10^−11^ (purple bars), alongside the number of fine-mapped variants from these loci with posterior inclusion probability PIP > 0.9 (pink bars).

**Supplementary Figure 6:**
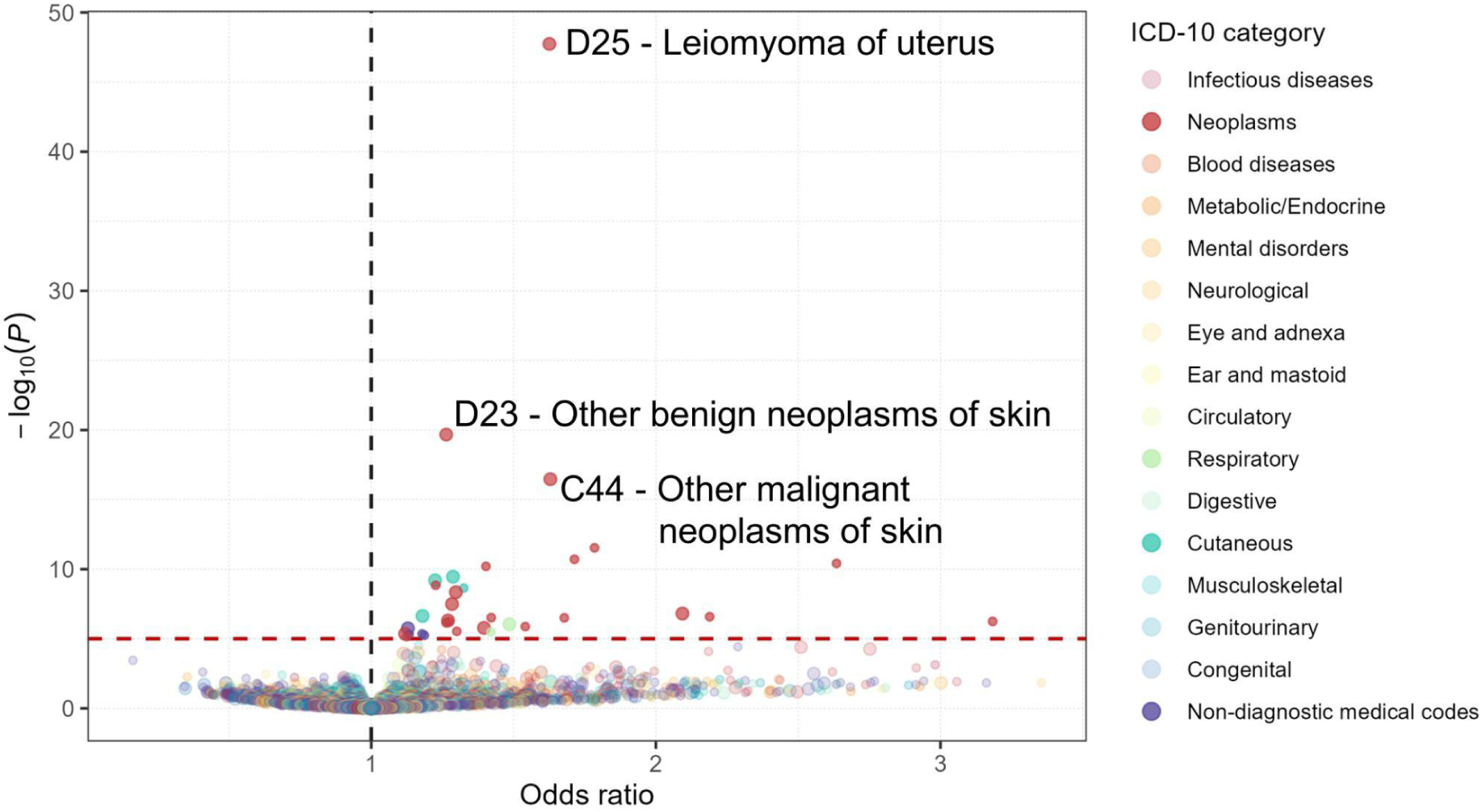
PheWAS of TP53 variant rs78378222. ICD-10-based phenome-wide association study of the TP53 3′UTR variant rs78378222, shown as a scatter plot of odds ratio versus −log10(P-value). Each point represents one ICD-10 trait, with colours indicating trait groups and point size distinguishing traits defined at the major or minor ICD-10 category level. Odds ratios and P-values were taken from GWAS results obtained using logistic regression. The red dashed horizontal line marks the Bonferroni significance threshold at P < 9.11 × 10^−6^. The three strongest associations by P-value are highlighted.

**Supplementary Figure 7:**
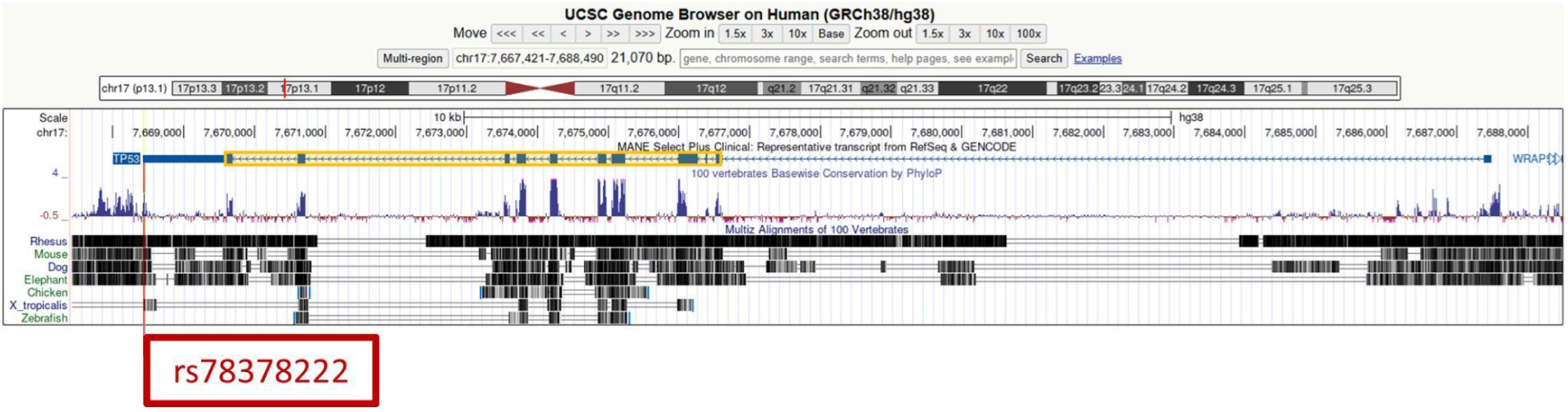
UCSC Genome Browser view of the TP53 locus on chromosome 17p13.1. The TP53 coding region is highlighted by the yellow box, and the 3′UTR variant rs78378222 is marked in red. The conservation track shows that the sequence surrounding the variant is highly conserved across mammals. Image obtained from the UCSC Genome Browser on 21st April 2026, https://genome-euro.ucsc.edu/cgi-bin/hgTracks?db=hg38.

**Supplementary Figure 8:**
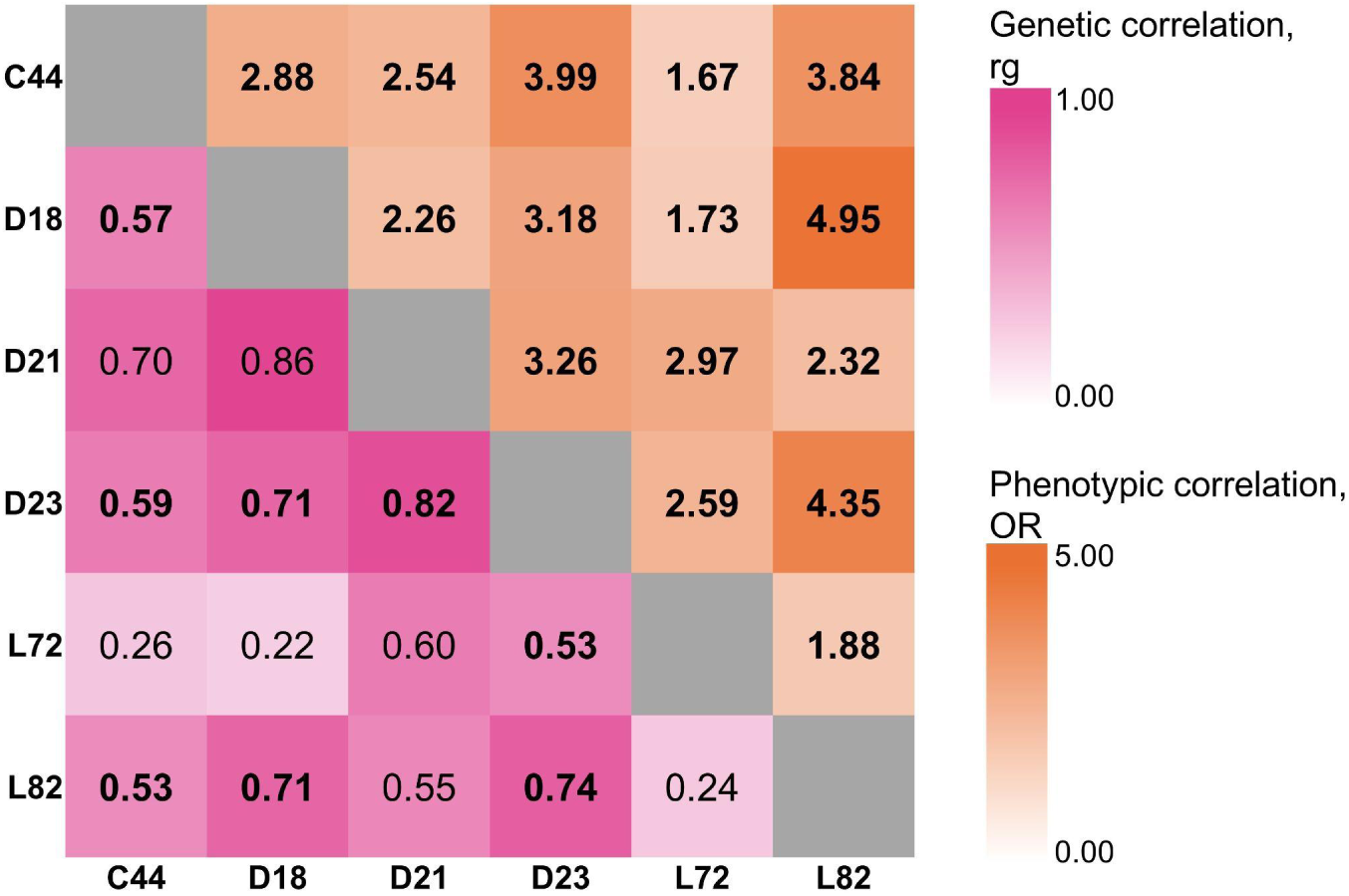
Genetic and phenotypic correlations across traits sharing the TP53 locus signal. Heatmap of pairwise relationships across six dermatology-related traits associated with the TP53 locus. The lower left corner shows genetic correlations estimated by LD score regression, coloured according to rg values, and the upper corner shows phenotypic correlations expressed as odds ratios obtained via logistic regression between trait carriers, coloured according to effect size. Traits included were C44, other malignant neoplasms of skin; D18, haemangioma and lymphangioma, any site; D21, other benign neoplasms of connective and other soft tissue; D23, other benign neoplasms of skin; L72, follicular cysts of skin and subcutaneous tissue; and L82, seborrhoeic keratosis. Values displayed in bold indicate pairwise relationships that remained significant after Bonferroni correction (P < 6.66 × 10^−6^). Grey diagonal cells indicate self-comparisons and were not evaluated.

**Supplementary Figure 9:**
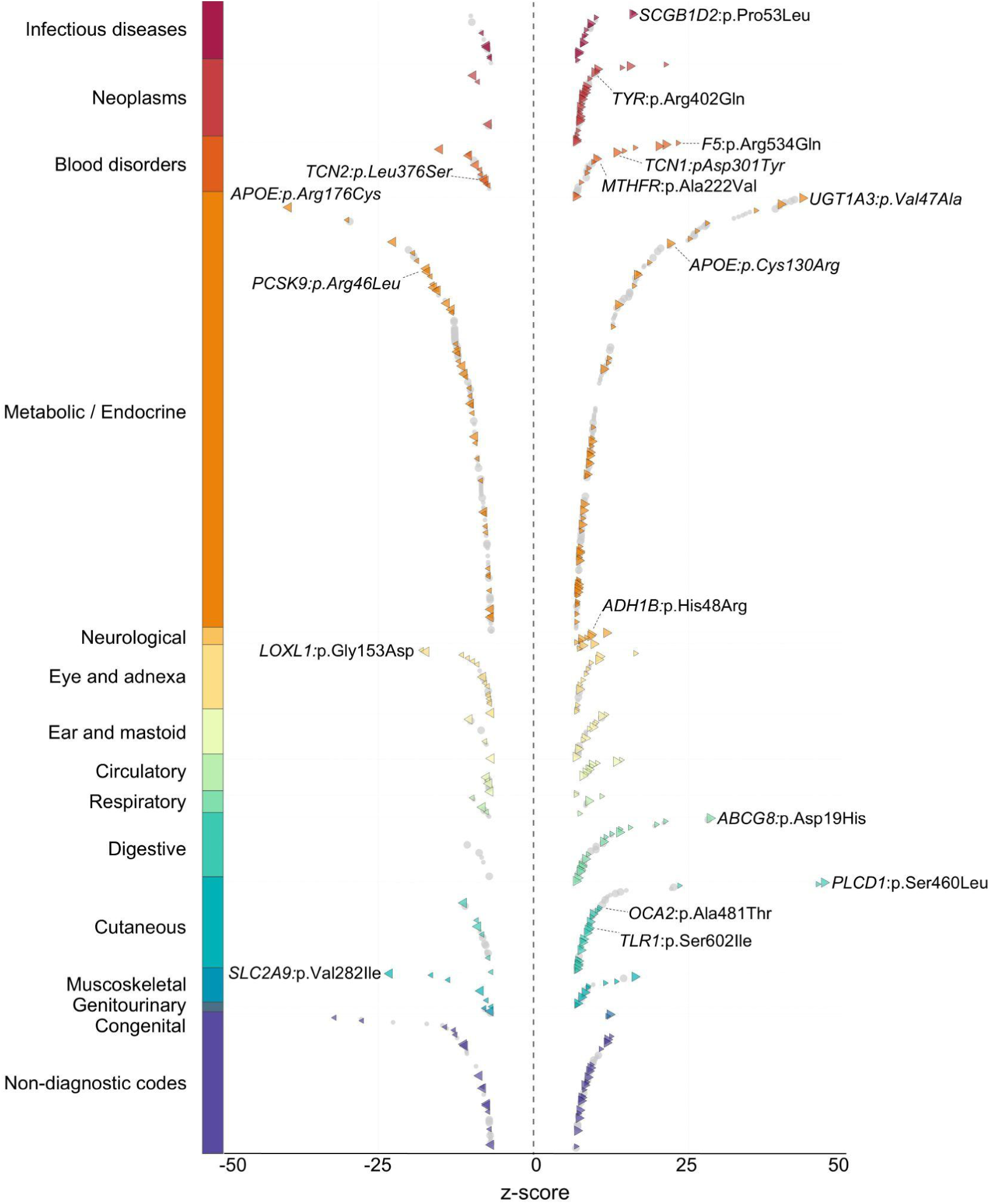
Disease-category distribution of lead coding variant associations. Scatter plot of 754 non-HLA coding variant-trait associations identified in EstBB, grouped by ICD-10 disease category. Each point represents one coding variant-trait pair and is positioned by ICD-10 category on the y-axis and association Z-score on the x-axis. Positive Z-scores indicate increased disease risk, whereas negative Z-scores indicate protective associations. Non-lead variants within each locus are shown in grey. Lead variants, defined as the strongest absolute Z-score association within a 2-Mb locus for each trait, are coloured by ICD-10 category. The dashed horizontal line marks Z = 0.

**Supplementary Figure 10:**
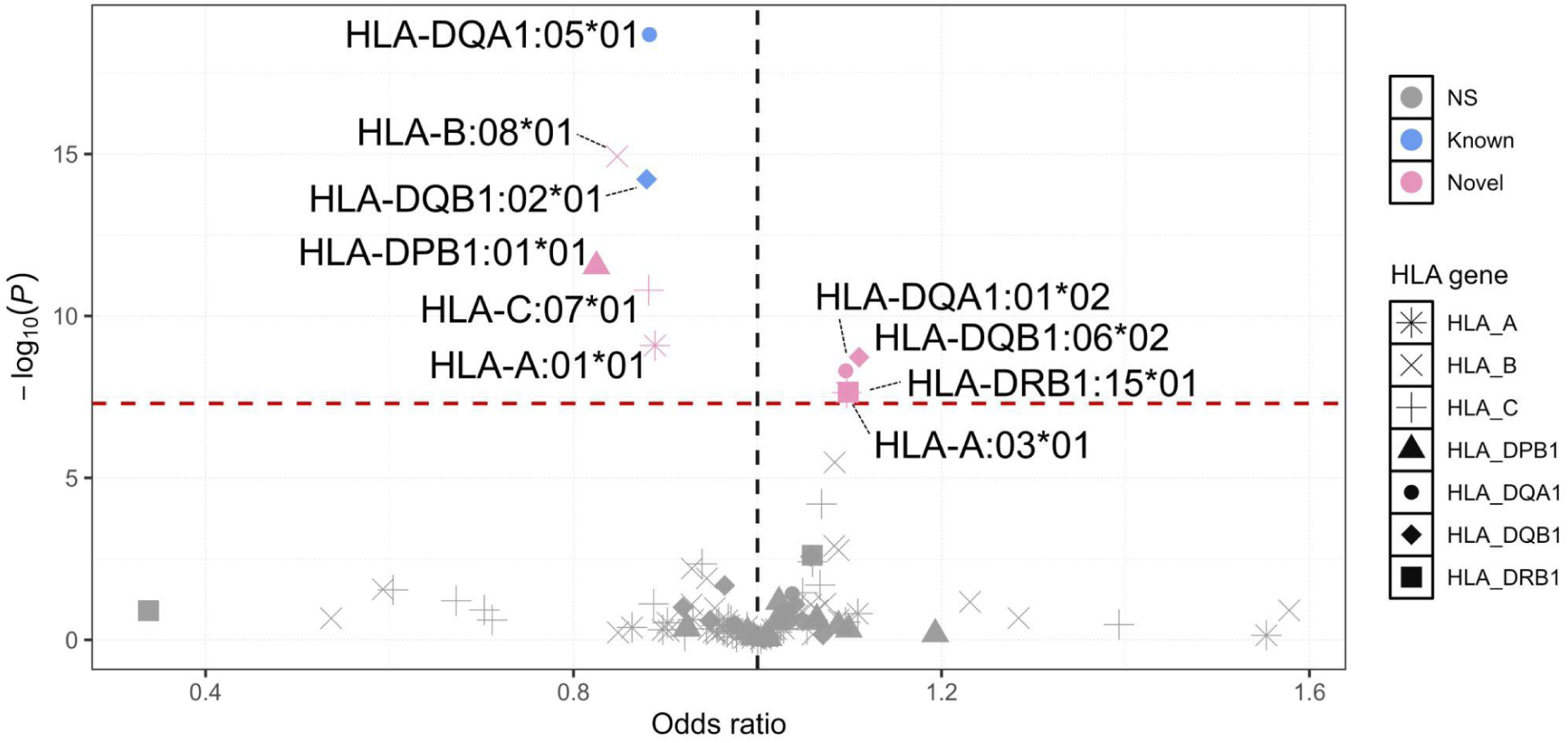
Association plot of HLA alleles in rosacea. Association results for HLA alleles in rosacea (ICD-10 code L71), shown as a scatter plot of odds ratio versus −log10(P-value). Each point represents one HLA allele. Point shape indicates the corresponding main HLA gene, and colours distinguish non-significant associations (gray), genome-wide significant known associations (blue), and genome-wide significant novel associations (pink). Odds ratios and P-values were obtained from logistic regression based association testing using REGENIE. The red dashed horizontal line marks the genome-wide significance threshold at −log10(P-value) = 7.3, corresponding to P < 5 × 10^−8^, and the vertical dashed line marks the null effect at odds ratio = 1. Only variants with INFO ≥ 0.9 are shown. NS = not significant.

**Supplementary Figure 11:**
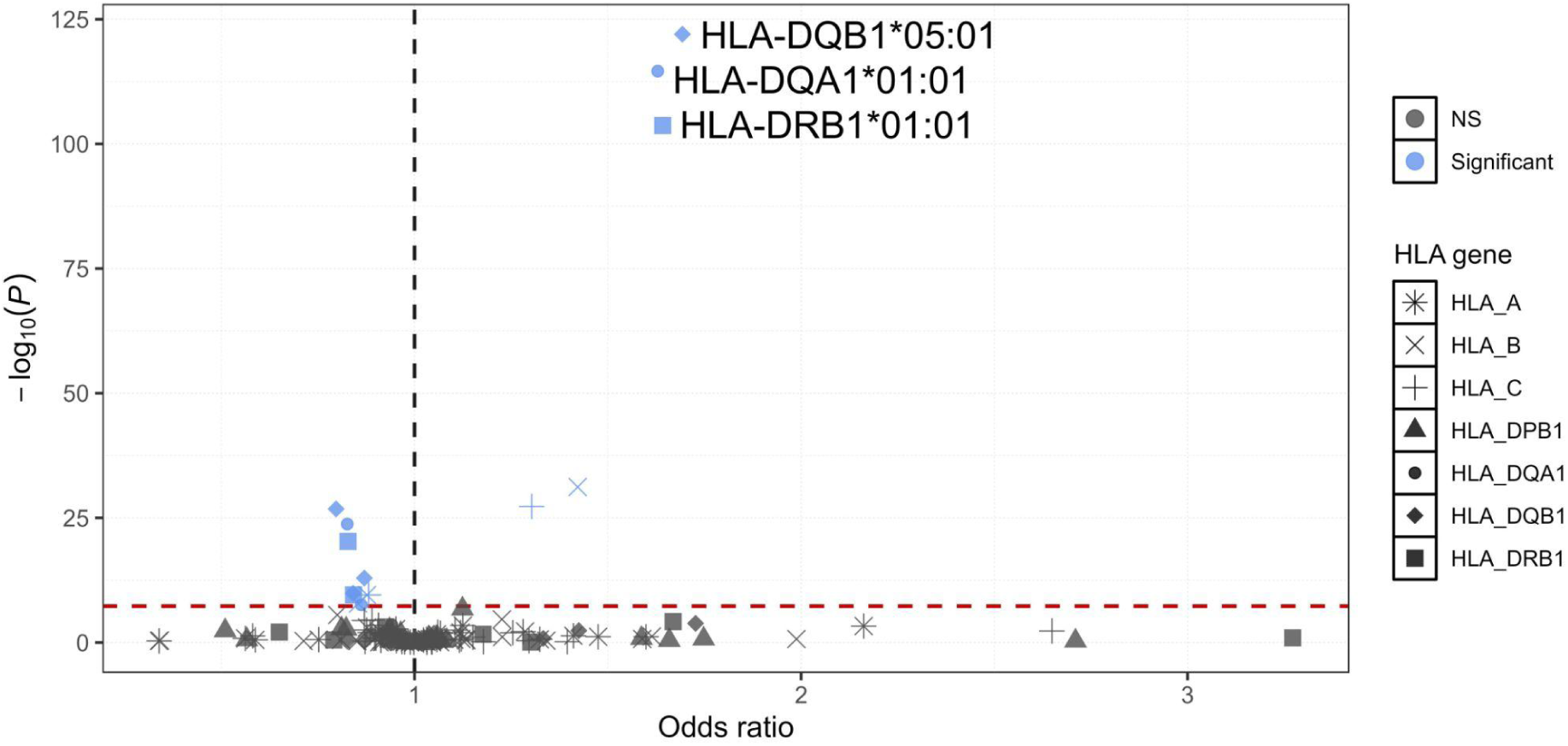
Association plot of HLA alleles in pityriasis versicolor. Association results for HLA alleles in pityriasis versicolor (ICD-10 code B36.0), shown as a scatter plot of odds ratio versus −log10(P-value). Each point represents one HLA allele. Point shape indicates the corresponding main HLA gene, and colours distinguish non-significant associations (gray), and genome-wide significant known associations (blue). Odds ratios and P-values were obtained from logistic regression based association testing using REGENIE. The red dashed horizontal line marks the genome-wide significance threshold at −log10(P-value) = 7.3, corresponding to P < 5 × 10^−8^, and the vertical dashed line marks the null effect at odds ratio = 1. Only variants with INFO ≥ 0.9 are shown. Top three variants by p-value significance are highlighted.

**Supplementary Figure 12:**
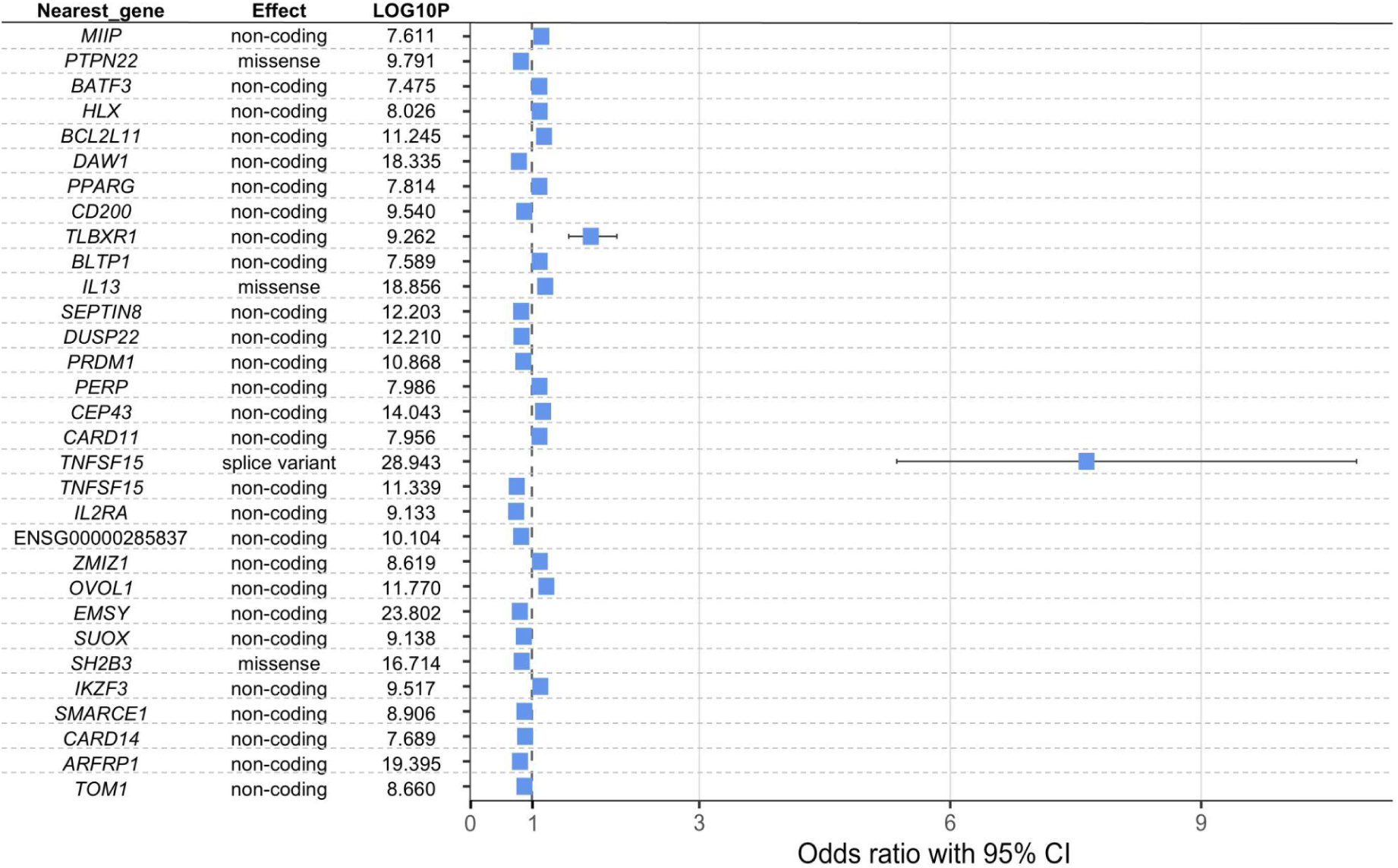
Pityriasis versicolor GWAS lead signals. Lead genome-wide association study signals for pityriasis versicolor (ICD-10 code B36.0), shown as odds ratios with 95% confidence intervals. Each row represents one lead variant, with the nearest gene, variant effect, and −log10(P) shown on the left. Squares indicate effect estimates and horizontal lines indicate 95% confidence intervals. The dashed vertical line marks an odds ratio of 1. The TNFSF15 splice variant showed the strongest effect estimate among the loci.

**Supplementary Figure 13:**
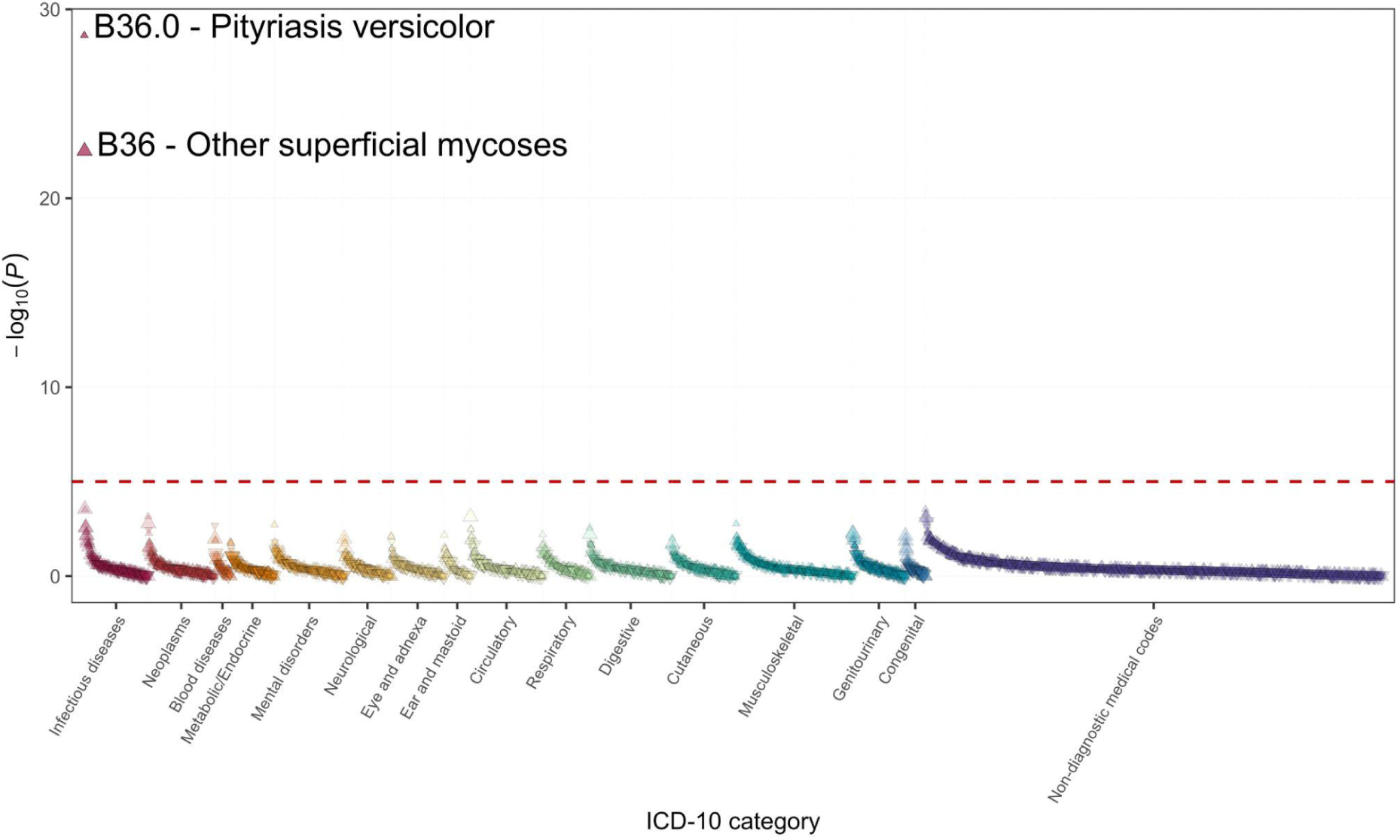
PheWAS of TNFSF15 variant rs1473488154. ICD-10-based phenome-wide association study of the TNFSF15 splice variant rs1473488154, shown as a category-based scatter plot of ICD-10 traits versus −log10(P-value). Each triangle represents one ICD-10 trait, coloured by trait group, with triangle size distinguishing traits defined at the major or minor ICD-10 category level. Triangle orientation indicates the direction of effect, corresponding to odds ratios above or below 1. P-values and odds ratios were taken from GWAS results obtained using logistic regression. The red dashed horizontal line marks the Bonferroni significance threshold at P < 9.11 × 10^−6^, and the two Bonferroni-significant associations are highlighted.

**Supplementary Figure 14:**
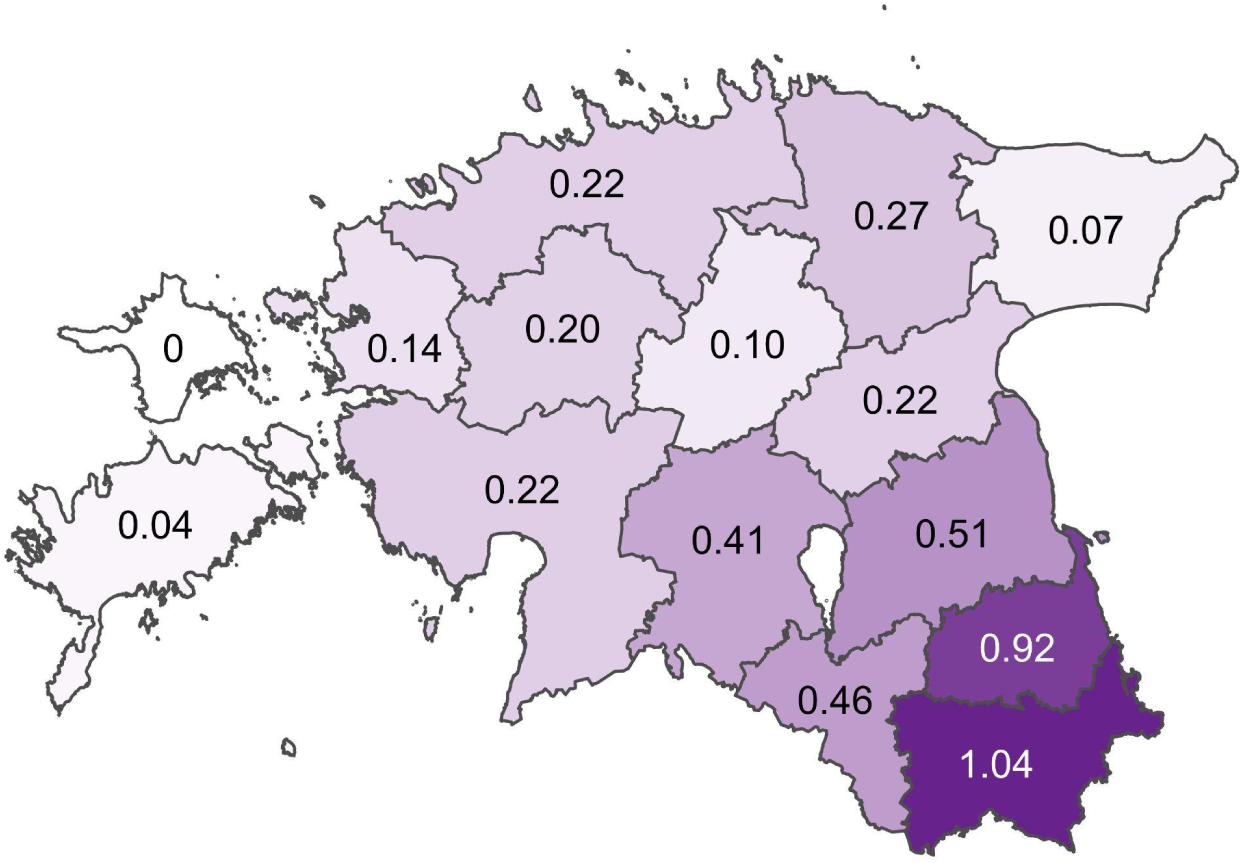
Geographical distribution of TNFSF15 splice variant carriers by birth county in Estonia. Numbers indicate the percentage of individuals carrying the variant within each birth county, and darker shading indicates a higher carrier proportion.

## Notes

### Competing Interest Statement

The authors have declared no competing interest.

### Author Declarations

The activities of the EstBB are regulated by the Human Genes Research Act, which was adopted in 2000 specifically for the operations of EstBB. Individual level data analysis in EstBB was carried out under ethical approval 1.1-12/624 from the Estonian Committee on Bioethics and Human Research (Estonian Ministry of Social Affairs), using data according to release application 3-10/GI/31689 from the Estonian Biobank. Study subjects in FinnGen provided informed consent for biobank research, based on the Finnish Biobank Act. Alternatively, separate research cohorts, collected prior the Finnish Biobank Act came into effect (in September 2013) and start of FinnGen (August 2017), were collected based on study-specific consents and later transferred to the Finnish biobanks after approval by Fimea (Finnish Medicines Agency), the National Supervisory Authority for Welfare and Health. Recruitment protocols followed the biobank protocols approved by Fimea. The Coordinating Ethics Committee of the Hospital District of Helsinki and Uusimaa (HUS) statement number for the FinnGen study is Nr HUS/990/2017. The FinnGen study is approved by Finnish Institute for Health and Welfare (permit numbers: THL/2031/6.02.00/2017, THL/1101/5.05.00/2017, THL/341/6.02.00/2018, THL/2222/6.02.00/2018, THL/283/6.02.00/2019, THL/1721/5.05.00/2019 and THL/1524/5.05.00/2020), Digital and population data service agency (permit numbers: VRK43431/2017-3, VRK/6909/2018-3, VRK/4415/2019-3), the Social Insurance Institution (permit numbers: KELA 58/522/2017, KELA 131/522/2018, KELA 70/522/2019, KELA 98/522/2019, KELA 134/522/2019, KELA 138/522/2019, KELA 2/522/2020, KELA 16/522/2020), Findata permit numbers THL/2364/14.02/2020, THL/4055/14.06.00/2020, THL/3433/14.06.00/2020, THL/4432/14.06/2020, THL/5189/14.06/2020, THL/5894/14.06.00/2020, THL/6619/14.06.00/2020, THL/209/14.06.00/2021, THL/688/14.06.00/2021, THL/1284/14.06.00/2021, THL/1965/14.06.00/2021, THL/5546/14.02.00/2020, THL/2658/14.06.00/2021, THL/4235/14.06.00/2021, Statistics Finland (permit numbers: TK-53-1041-17 and TK/143/07.03.00/2020 (earlier TK-53-90-20) TK/1735/07.03.00/2021, TK/3112/07.03.00/2021) and Finnish Registry for Kidney Diseases permission/extract from the meeting minutes on 4th July 2019. The Biobank Access Decisions for FinnGen samples and data utilized in FinnGen Data Freeze 12 include: THL Biobank BB2017_55, BB2017_111, BB2018_19, BB_2018_34, BB_2018_67, BB2018_71, BB2019_7, BB2019_8, BB2019_26, BB2020_1, BB2021_65, Finnish Red Cross Blood Service Biobank 7.12.2017, Helsinki Biobank HUS/359/2017, HUS/248/2020, HUS/430/2021 paragraph28, paragraph29, HUS/150/2022 paragraph12, paragraph13, paragraph14, paragraph15, paragraph16, paragraph17, paragraph18, paragraph23, paragraph58, paragraph59, HUS/128/2023 paragraph18, Auria Biobank AB17-5154 and amendment #1 (August 17 2020) and amendments BB_2021-0140, BB_2021-0156 (August 26 2021, Feb 2 2022), BB_2021-0169, BB_2021-0179, BB_2021-0161, AB20-5926 and amendment #1 (April 23 2020) and it's modifications (Sep 22 2021), BB_2022-0262, BB_2022-0256, Biobank Borealis of Northern Finland_2017_1013, 2021_5010, 2021_5010 Amendment, 2021_5018, 2021_5018 Amendment, 2021_5015, 2021_5015 Amendment, 2021_5015 Amendment_2, 2021_5023, 2021_5023 Amendment, 2021_5023 Amendment_2, 2021_5017, 2021_5017 Amendment, 2022_6001, 2022_6001 Amendment, 2022_6006 Amendment, 2022_6006 Amendment, 2022_6006 Amendment_2, BB22-0067, 2022_0262, 2022_0262 Amendment, Biobank of Eastern Finland 1186/2018 and amendment 22paragraph/2020, 53paragraph/2021, 13paragraph/2022, 14paragraph/2022, 15paragraph/2022, 27paragraph/2022, 28paragraph/2022, 29paragraph/2022, 33paragraph/2022, 35paragraph/2022, 36paragraph/2022, 37paragraph/2022, 39paragraph/2022, 7paragraph/2023, 32paragraph/2023, 33paragraph/2023, 34paragraph/2023, 35paragraph/2023, 36paragraph/2023, 37paragraph/2023, 38paragraph/2023, 39paragraph/2023, 40paragraph/2023, 41paragraph/2023, Finnish Clinical Biobank Tampere MH0004 and amendments (21.02.2020 & 06.10.2020), BB2021-0140 8paragraph/2021, 9paragraph/2021, paragraph9/2022, paragraph10/2022, paragraph12/2022, 13paragraph/2022, paragraph20/2022, paragraph21/2022, paragraph22/2022, paragraph23/2022, 28paragraph/2022, 29paragraph/2022, 30paragraph/2022, 31paragraph/2022, 32paragraph/2022, 38paragraph/2022, 40paragraph/2022, 42paragraph/2022, 1paragraph/2023, Central Finland Biobank 1-2017, BB_2021-0161, BB_2021-0169, BB_2021-0179, BB_2021-0170, BB_2022-0256, BB_2022-0262, BB22-0067, Decision allowing to continue data processing until 31st Aug 2024 for projects: BB_2021-0179, BB22-0067,BB_2022-0262, BB_2021-0170, BB_2021-0164, BB_2021-0161, and BB_2021-0169, and Terveystalo Biobank STB 2018001 and amendment 25th Aug 2020, Finnish Hematological Registry and Clinical Biobank decision 18th June 2021, Arctic biobank P0844: ARC_2021_1001.

### Summary of Updates

The manuscript has been extensively revised and substantially expanded since the previous version. Major updates include reorganising the manuscript structure, revising the introduction and discussion, implementing new analyses, and clarifying methodological details throughout. Both main and supplementary figures, as well as supplementary tables, were comprehensively updated, with several panels replaced, redesigned, or newly added to improve clarity and data presentation. Results were reevaluated and interpreted in light of the expanded analyses, leading to revised conclusions and improved contextualisation within the current literature. We also corrected inconsistencies and updated references.

